# Genome-wide association study between SARS-CoV-2 single nucleotide polymorphisms and virus copies during infections

**DOI:** 10.1101/2024.03.02.24303655

**Authors:** Ke Li, Chrispin Chaguza, Julian Stamp, Yi Ting Chew, Nicholas F.G. Chen, David Ferguson, Sameer Pandya, Nick Kerantzas, Wade Schulz, Yale SARS-CoV-2 Genomic Surveillance Initiative, Anne M. Hahn, C Brandon Ogbunugafor, Virginia E. Pitzer, Lorin Crawford, Daniel M. Weinberger, Nathan D. Grubaugh

**Author notes:** Authors listed at the end of the manuscript.

## Abstract

Significant variations have been observed in viral copies generated during SARS-CoV-2 infections. However, the factors that impact viral copies and infection dynamics are not fully understood, and may be inherently dependent upon different viral and host factors. Here, we conducted virus whole genome sequencing and measured viral copies using RT-qPCR from 9,902 SARS-CoV-2 infections over a 2-year period to examine the impact of virus genetic variation on changes in viral copies adjusted for host age and vaccination status. Using a genome-wide association study (GWAS) approach, we identified multiple single-nucleotide polymorphisms (SNPs) corresponding to amino acid changes in the SARS-CoV-2 genome associated with variations in viral copies. We further applied a marginal epistasis test to detect interactions among SNPs and identified multiple pairs of substitutions located in the spike gene that have non-linear effects on viral copies. We also analyzed the temporal patterns and found that SNPs associated with increased viral copies were predominantly observed in Delta and Omicron BA.2/BA.4/BA.5/XBB infections, whereas those associated with decreased viral copies were only observed in infections with Omicron BA.1 variants. Our work showcases how GWAS can be a useful tool for probing phenotypes related to SNPs in viral genomes that are worth further exploration. We argue that this approach can be used more broadly across pathogens to characterize emerging variants and monitor therapeutic interventions.

**Author Summary:** Our study explores why viral load (copies measured by RT-qPCR) varies during SARS-CoV-2 infections by analyzing viral mutations and measuring viral copies in 9,902 individuals over two years. We aimed to understand how genetic differences in SARS-CoV-2 influence viral copies, considering host age and vaccination status. Using a genome-wide association study (GWAS), we identified several single-nucleotide polymorphisms (SNPs) in the virus linked to variations in viral levels. Notably, interactions between certain SNPs in the spike gene had non-linear effects on viral copies. Our analysis revealed that SNPs associated with higher viral copies were common in Delta and Omicron BA.2/BA.4/BA.5/XBB variants, while those linked to lower levels were mainly found in Omicron BA.1. This research highlights GWAS as a powerful tool for exploring virus genetics and suggests it can be broadly applied to monitor new variants of COVID-19 and other infectious diseases.

## Introduction

Continued SARS-CoV-2 transmission and evolution has propelled the COVID-19 pandemic. Peak viral replication in the upper respiratory tract occurs during the first few days of infection [1]. The viral load (or copies measured by RT-qPCR) in patient samples are valuable data to understand infection dynamics, such as inferring the likelihood of disease transmission [2]. However, it is challenging to use viral load data, and the challenge often arises from significant variations in viral load dynamics among sampled cases, which can be associated with 1) host heterogeneity, e.g., age [3] and vaccination status [6–8]; 2) distinct inherent properties of virus variants or sublineages [9], and 3) different sampling times [10]. For example, sampling during the early stages of infection may yield higher viral loads compared to later stages after viral replication has reached its peak. Nevertheless, the relative importance of these factors influencing viral load has not been completely explored [11,12].

Genome-wide association studies (GWAS) have emerged as a useful tool in the field of genetics, providing an approach to unraveling the complex interplay between genetic variations and observable traits, including diseases and drug resistance, as reviewed in [13]. Several studies have employed GWAS analysis to identify and investigate the association between human genetic variations across different individuals and the severity of COVID-19, shedding light on genetic variations that are related to severe infections [15–17]. However, few studies have utilized a GWAS method to study associations between the viral genome and viral traits [19–22]. The confluence of the extensive existing research on SARS-CoV-2 mutations and the millions of infections that have been sequenced provides us the opportunity to evaluate the application of GWAS for viral genomics. The hypothesis-free approach has the potential to enhance our understanding of genetic determinants influencing viral fitness and evolution and further inform effective public health strategies aimed at mitigating the spread and impact of SARS-CoV-2.

In this work, we aim to investigate the impact of intrinsic viral genetic substitutions (i.e., single nucleotide polymorphisms [SNPs]) on the changes in viral copies, adjusted for host age and vaccination status. For this, we apply a viral GWAS analysis to SARS-CoV-2 genomic sequencing and standardized RT-qPCR data collected from the Yale New Haven Hospital from February 2021 to March 2023. Using whole genome sequencing data on SARS-CoV-2 infections, along with relevant laboratory and patient metadata, we identify associations between viral SNPs and viral copies for different variants of concern (VOCs). We then examine the temporal pattern of identified SNPs by constructing a phylogenetic tree, drawing upon subsamples, and analyzing the time series of the fraction of SNPs occurring in the sequences. This multifaceted analysis contributes to unraveling the complex dynamics of SARS-CoV-2 infections, providing valuable insights into the underlying viral SNPs that influence viral copies in different VOCs.

## Results

### Viral copies vary in SARS-CoV-2 infections

To better understand how SARS-CoV-2 viral load varies in infected individuals, we analyzed the viral copy data, along with associated host metadata (i.e., age and vaccination status), and genome sequencing data from a cohort of patients tested at the Yale New Haven Hospital (YNHH) located in Connecticut, US. We selected 9902 whole genome sequences with available viral copy data generated from remnant SARS-CoV-2 diagnostic samples over a 2-year period, from 03-Feb-2021 to 21-Mar-2023 (**Fig. 1A).** The VOCs that we identified in our dataset during the sampling period included Alpha (n = 809), Delta (n = 1278), Gamma (n = 36), BA.1 (n = 1818), BA.2 (n = 2432), BA.4 (n = 293), BA.5 (n = 1992), XBB (n = 698), and the pre-VOC variant (named ‘Other’, n = 546). We conducted RT-qPCR using a standardized assay targeting the nucleocapsid (CDC ‘N1’ primers) for each sample to allow for cross-sample comparisons [23], except for a period during October 2021 when the PCR data were not generated. Across all samples, the viral copies, expressed as log10(viral copies per milliliter (Genome Equivalents/ml)), exhibited variations, ranging from 3.60 to 10.55, with a median value of 7.26 (**Fig. 1B**). The variations in viral copies could be attributed either to the introduction and/or replacement of different VOCs, each with its own epidemic curve, or to the stochasticity from the sampling process. To reduce stochastic effects, we aggregated the viral copies by month and still observed large variations in the viral copies across the months, albeit with no consistent trend (**Fig. 1C**). Notably, we observed the lowest median value of viral copies (median = 6.49) in February 2022, during which 96.3% of the sampled sequences tested positive for BA.1 infections. By contrast, we observed the highest median value of viral copies (median = 7.70) in June 2022, during which the sampled sequences tested positive for BA.2 (64.9%), BA.4 (6%), or BA.5 (29.1%) infections. Taken together, we showed a wide range of viral copies in the sampled SARS-CoV-2 infections with different VOCs, utilizing data from genomic surveillance and standardized RT-qPCR tests.

**Figure 1.**
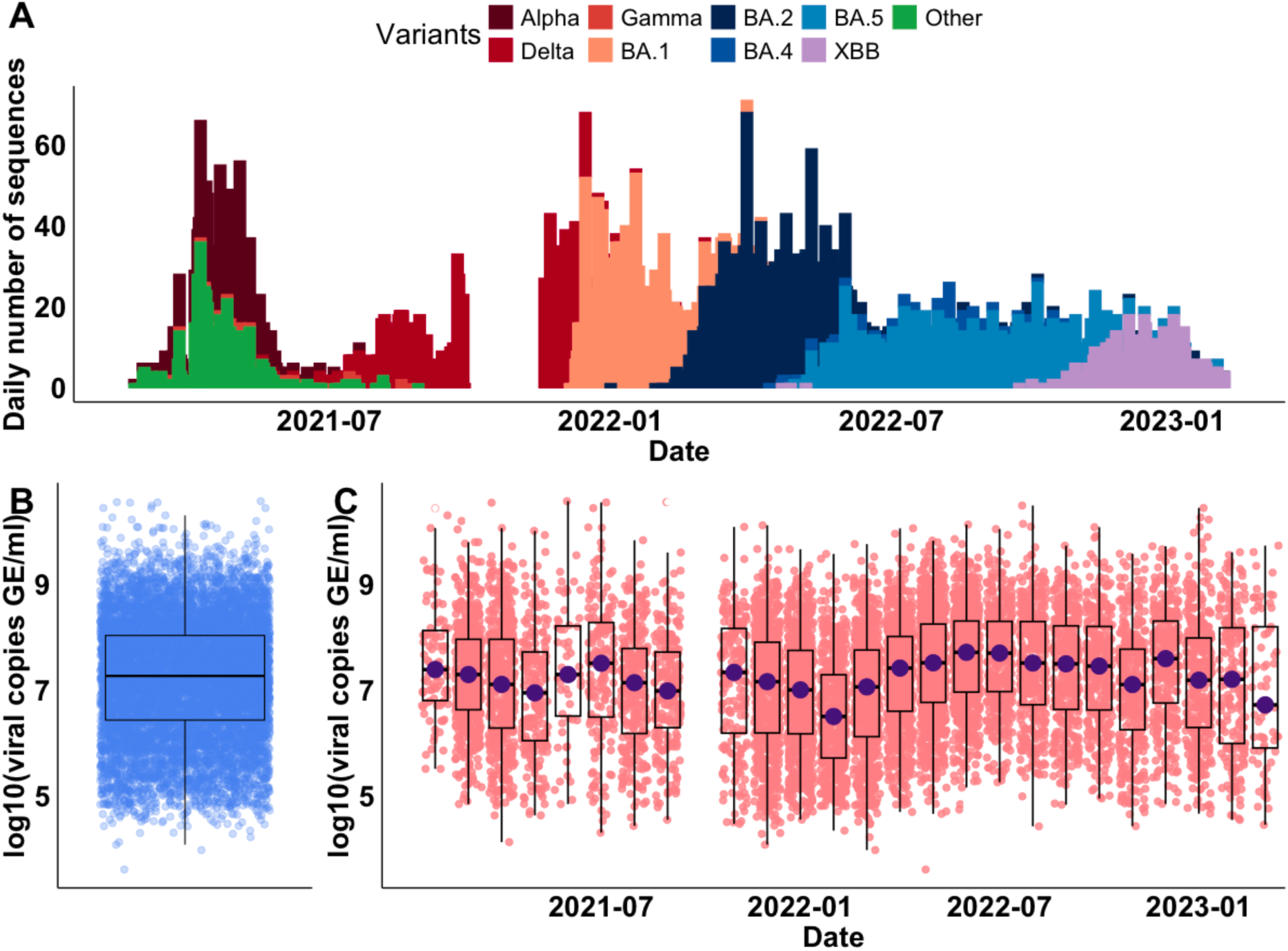
Genomic sequences of SARS-CoV-2 infections and associated viral copies from cross-sectional samples collected in Connecticut, US. (**A**) The daily number of genomic sequences of SARS-CoV-2 VOCs from February 2021 to March 2023. (**B**) The summary of viral copies of all samples, expressed as *log*10(viral copies per milliliter). (**C**) The summary of viral copies aggregated by month. The data gap in October 2021 is because we were unable to conduct PCR to obtain viral copies during this time.

### Viral copies correlate with age and variants, but not with vaccination status

Having uncovered a large variability in the observed viral copies from the samples, we next assessed the factors associated with these changes. To do this, we first summarized and compared viral copies in various age groups (**Fig. 2A**). A positive correlation has been previously reported between age and SARS-CoV-2 viral copies, showing that younger age groups had lower viral copies independent of gender and/or symptom duration [24]. We observed a similar result in our dataset and found that the oldest age group (i.e., >70 years old) had the highest viral copies compared with other age groups (mean = 7.47, 95% confidence interval (CI): [5.12, 9.49], *p* < 0.001, Wilcoxon signed-rank test). For the effect of vaccination on viral copies, some studies have demonstrated that although vaccination reduced the risk of infections with the Delta variant, no significant difference in peak viral copies was found between fully vaccinated and unvaccinated individuals [6,7,25]. In contrast, other studies have shown that vaccination reduced viral copies in BA.1 infections among boosted individuals compared to unvaccinated ones [8]. These results suggest the effect of vaccination on viral copies may depend on the characteristics of the infecting SARS-CoV-2 variant. We compared viral copies among groups with different vaccination statuses to assess the impact of vaccination on viral copies (**Fig. 2B**), and no statistically significant differences were detected between the groups in our data (*p* > 0.05, Wilcoxon signed-rank test). Finally, we compared viral copies stratified by variant category (**Fig. 2C**). Combining samples collected from all age and vaccination status groups for each variant, we found that the overall mean values of viral copies were lowest for infections with BA.1 (mean = 6.83, 95% CI: [4.87, 8.87], *p* < 0.001, Wilcoxon signed-rank test) compared to infections with other all non-BA.1 variants.

**Figure 2.**
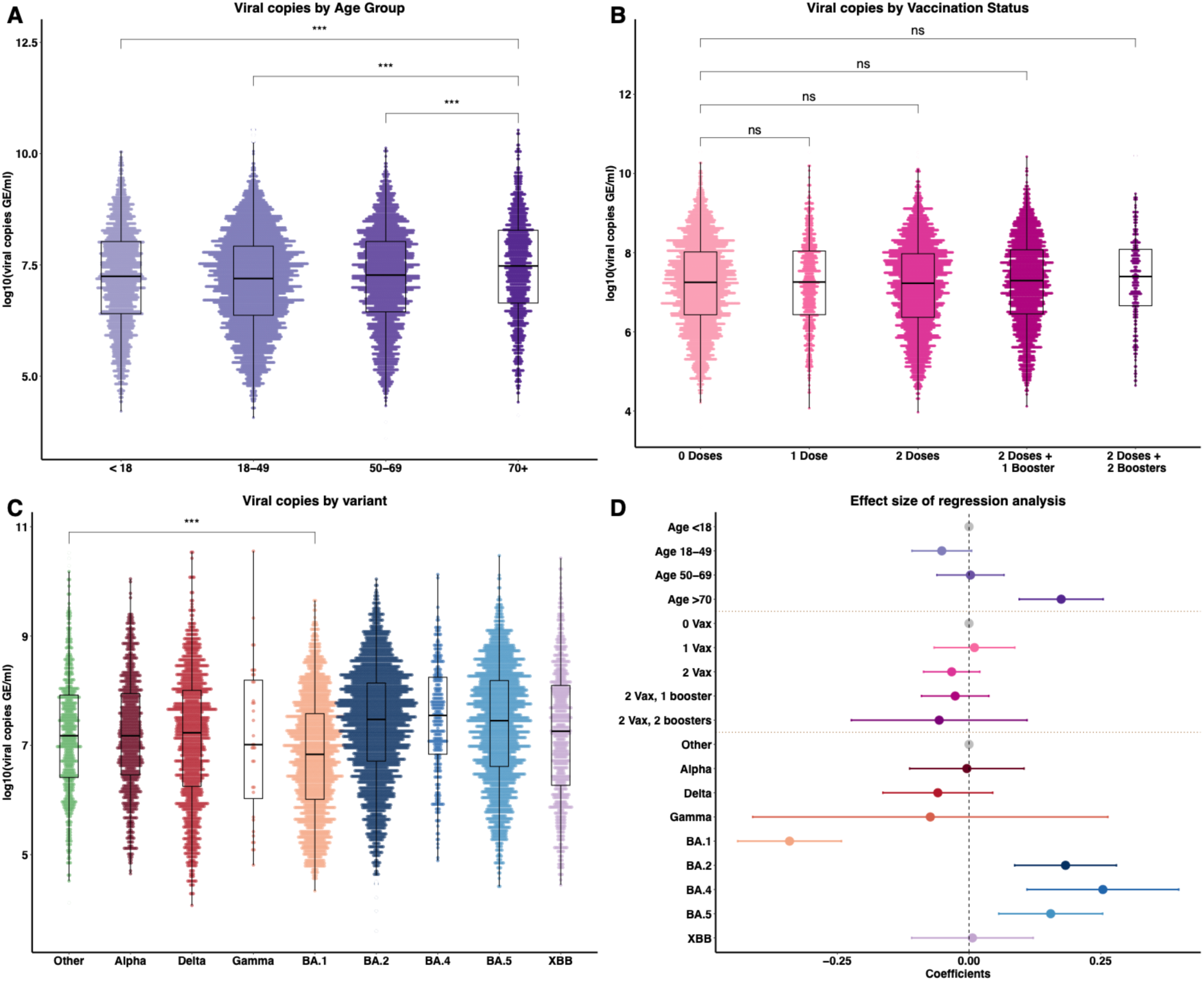
Viral copies by category and regression analysis results. Comparison of viral copies stratified by (**A**) age groups, (**B**) vaccination statuses, (**C**) variant of concerns. (**D**) Association of age, vaccination status, and VOCs with viral copies, expressed as *log*_10_(viral copies per milliliter (Genome Equivalents/ml)). The reference groups (in gray) are Age <18 years old, 0 doses of vaccination, and the Other variant, respectively. The positive coefficients indicate the covariate is associated with higher viral copies value compared to the reference group, and vice versa. 0 Vax, 1 Vax, 2 Vax, 2 Vax 1 booster, and 2 vax 2 boosters denote vaccination statuses of 0 doses, 1 dose, 2 doses, 2 doses, and 1 booster, and 2 doses and 2 boosters, respectively, corresponding to the labels in (**B**). Results are shown as means with 95% confidence intervals. *** *p* < 0.001.

Since several factors may simultaneously impact the SARS-CoV-2 viral load, next, we sought to quantify the combined impact of age, vaccination status, and VOCs on the observed viral copies. To achieve this, we fitted a multivariate linear regression model, with viral copies as the outcome variable and age, vaccination, and VOCs as covariates (**Fig. 2D**). We found that the older age group (i.e., age >70 years old) had a positive association with viral copies (mean = 0.17, 95% CI: [0.09, 0.25], *p* < 0.001) compared with the reference group (i.e., age <18 years old). We also found that vaccination status was not associated with viral copies (i.e., 95% CIs of the vaccination coefficients span 0, *p* > 0.05). Notably, we showed that infections with BA.1 were associated with reduced viral copies, with a mean effect size of -0.34 (95% CI: [-0.44, -0.24], *p* < 0.001) in the same age group and vaccination status, compared to the Other variant. We also showed that infections with BA.2 (mean = 0.19, 95% CI: [0.09, 0.28], *p* < 0.001), BA.4, or BA.5 (mean = 0.17, 95% CI: [0.07, 0.26], *p* < 0.001) were associated with increased viral copies. Among them, infections with BA.4 were associated with the largest positive effect size (mean = 0.27, 95% CI: [0.12, 0.41], *p* < 0.001). Our findings demonstrated that variations in viral copies were associated with infections caused by different SARS-CoV-2 variants and the older age group. This implies that intrinsic factors of the viruses, such as genetic mutations among distinct VOCs, are key determinants impacting viral copies.

### Viral GWAS reveals SARS-CoV-2 SNPs associated with viral copies

Having demonstrated that changes in SARS-CoV-2 viral copies are associated with infections caused by different viral variants or strains, especially Omicron BA.1/BA.2/BA.4/BA.5 variants (**Fig. 2D**), we then sought to identify potential genetic mutations—specifically, SNPs—that contributed to these changes in viral copies. For this, we performed a GWAS analysis using high-quality genome sequences (i.e., genome coverage > 95%). We conducted whole-genome sequencing on the 9902 SARS-CoV-2 positive specimens collected from February 2021 to March 2023. Firstly, using Wuhan-Hu-1 (GenBank MN908937.3) as the reference genome, we identified 10,697 SNPs for further testing associated with viral copies as covariates. We then checked for the population structure of the 9902 genome sequences using a multidimensional scaling (MDS) method [26] (**Fig. S1**). We observed that Delta was an outgroup to other pre-Omicron variants (i.e., pre-VOC variant (Other), Alpha, and Gamma), and BA.1 was an outgroup to the BA.2/BA.4/BA.5/XBB cluster. In our model, we included the inferred four clusters based on the MDS-computed distance to capture the viral population structure. Clusters were defined using a k-means clustering method (**Fig. S1**). The host ages and vaccination status were also included in the model as covariates.

Using the linear regression model on viral copies for each SNP, adjusted for viral population structure and host factors, we identified 31 SNPs exceeding the permuted threshold for genome-wide significance (*p* = 4.67 × 10^-6^, dashed line, **Fig. 3A**). The threshold value was calculated as 0.05 divided by 10,697 SNPs [27]. We found that the observed distribution of *p*-values closely matches the expected distribution under the null hypothesis of no association (**Fig. S2A**). To ascertain whether those SNPs have a negative or positive impact on viral copies and evaluate their effect size, we extracted the coefficients (*β*) of the SNPs with *p* < 1 × 10^-10^ and their standard deviations (*σ*) from the regression model (dashed box, **Fig. 3B**). We then annotated the SNPs to identify the associated amino acids, and among them, 14 SNPs were non-synonymous (i.e., changed the amino acid; **Fig. 3C**). We found that a non-synonymous change N:R203M, located on the N gene, had the most significant association with increased viral copies (*p* = 2.68 × 10^-22^, *β* = 1.65, *σ* = 0.16). By contrast, the amino acid change most strongly associated with a negative effect on viral copies was ORF1ab:L5086I (*p* = 9.20 × 10^-20^, *β* = -1.20, *σ* = 0.13). We further conducted a marginal epistasis test [28–30] to detect the epistatic effects of SNPs on viral copies. We discovered multiple pairs of SNPs that exhibit positive epistatic effects on viral copies, with most interactions occurring in the S gene (**Fig. S3**).

**Figure 3.**
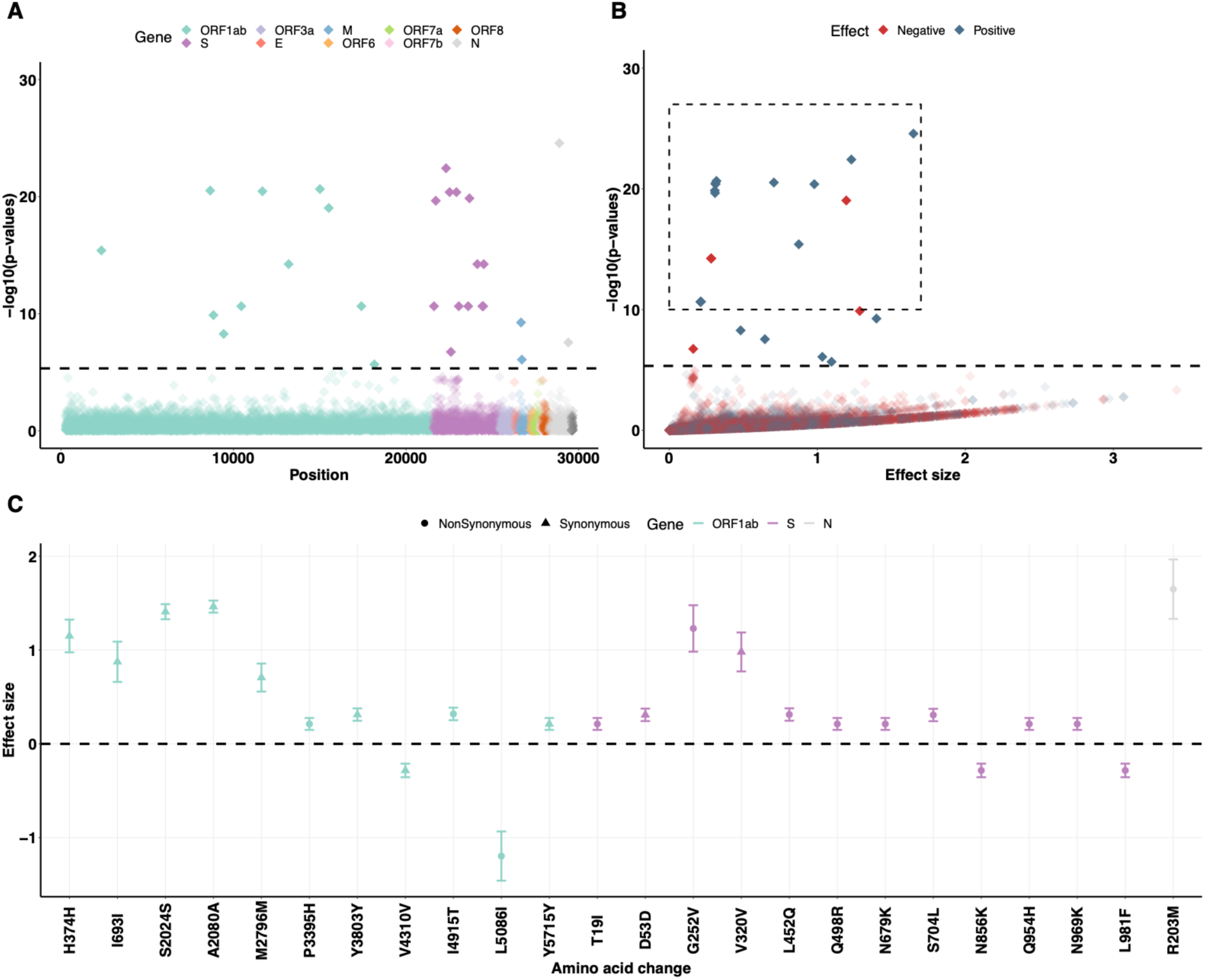
GWAS analysis identifies several single nucleotide polymorphisms (SNPs) that are associated with the changes in viral copies. (A) Genome-wide association results of the impact of identified SNPs on viral copies during SARS-CoV-2 infection. The dashed line indicates the permuted threshold for genome-wide significance *p* = 4.67 × 10^-5^ (0.05/10697 SNPs). Significant SNPs are shown with solid colors. (B) SNPs (with *p* < 1 × 10^-10^) that have positive (blue) or negative (red) effects on viral copies. (C) The corresponding synonymous (triangles) and non-synonymous (circles) amino acid changes that associate with increased or decreased viral copies. Data shown as means with 95% confidence intervals. The estimated effective sizes and associated standard deviations are given Table S1.

To assess the impact of adjusting for the population structure of the SARS-CoV-2 strains using the MDS components on the regression results, we conducted a sensitivity analysis on the genome sequences using the inferred MDS components from the pairwise SNP distance matrix of SARS-CoV-2 sequences as covariates. By doing this, we identified 113 SNPs exceeding the permuted threshold (**Fig. S4**). The observed distribution of *p*-values also closely matched the expected distribution under the null hypothesis of no association (**Fig. S2B**). The results may be more likely to reflect the SNPs that influence the viral copies dependent on lineage. We also examined the association between viral copies and SNPs after adjusting for the population structure based on the VOCs themselves, which broadly correspond to the identified sequence clusters. We showed that only a few SNPs were found (Figs. S5-8), mostly within the Omicron BA.2/BA.4/BA.5/XBB cluster (**Fig. S8**).

### The impact of amino acid changes on viral copies is dependent on the variant

Having identified the 14 non-synonymous SNPs with statistically significant effects on viral copies in our primary analysis, we next sought to understand the temporal patterns of the emergence of these amino acid changes (**Fig. 4**). To investigate the clustering of these SNPs, we randomly sampled approximately 120 genome sequences from each VOC category (only 36 sequences were available for Gamma in our dataset) and generated a phylogenetic tree drawing upon the subsamples (**Fig. 4A**). We found a clear pattern in how these mutations emerged by VOC (**Fig. 4A** heatmap). We found that all amino acid changes associated with a positive effect on viral copies were found in Delta and Omicron BA.2/BA.4/BA.5/XBB infections. Often, more than one amino acid change was observed in each sampled sequence, suggesting genetic linkage between these SNPs, as also shown in the epistasis test (**Fig. S3**), such as S:Q954H and N969K. In particular, we identified that the amino acid changes S:L452Q (*p* = 3.91 × 10^-25^, *β* = 0.34, *σ* = 0.03) and S704L (*p* = 1.35 × 10^-24^, *β* = 0.34, *σ* = 0.03) associated with a positive effect on viral copies were typically observed in combination with BA.2 infections—specifically, lineage BA.2.12.1. We also observed that the amino acid changes with negative effects on viral copies (ORF1ab:L5086I, S:N856K and L981F) were only associated with BA.1 infections.

**Figure 4.**
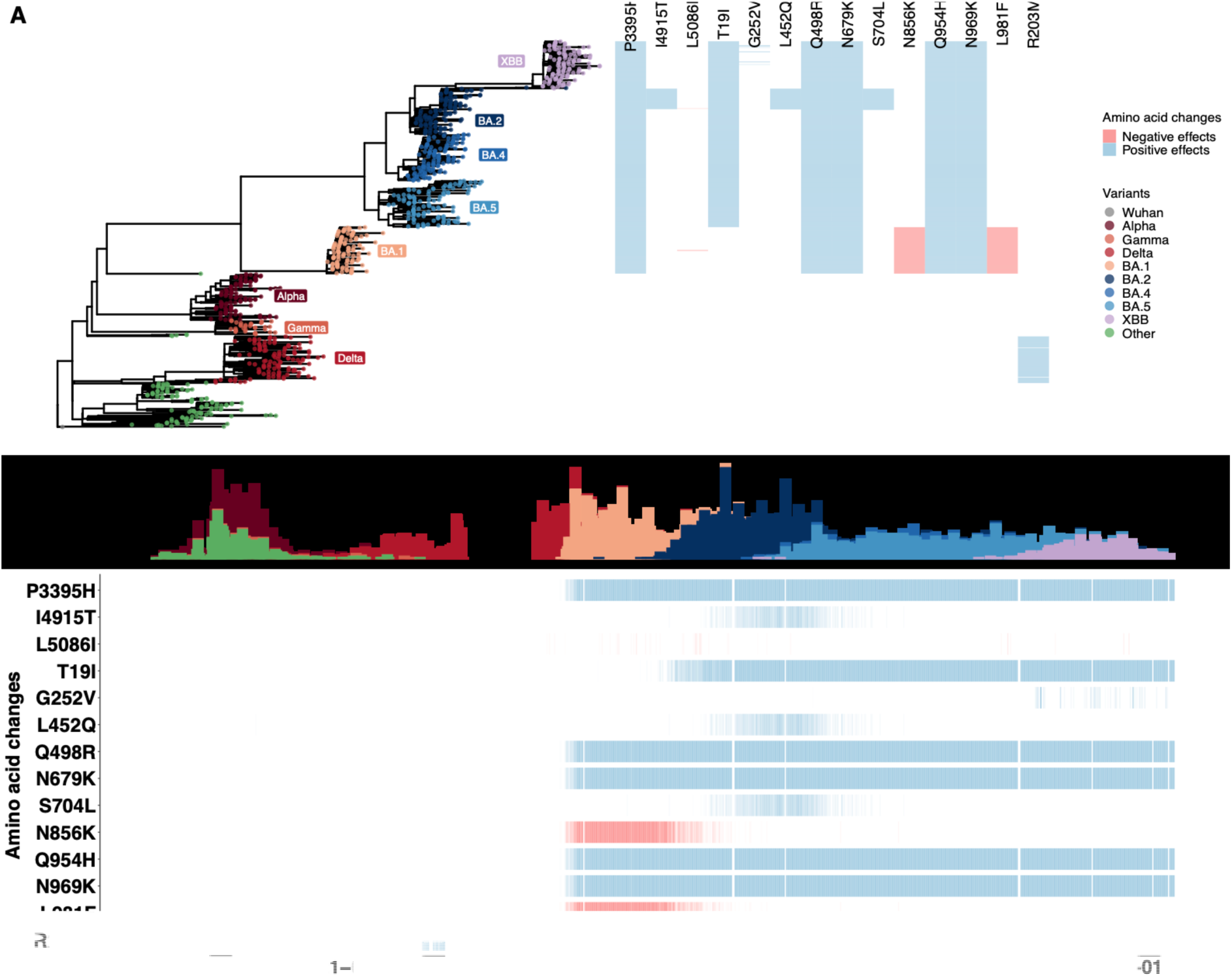
The temporal dynamics of non-synonymous amino acid changes in the ORF1ab gene (P3395H, I4915T and L5086I), S gene (T19I, G252V, L452Q, Q498R, N679K, S704L, N856K, Q954H, N969K and L981F), and N gene (R203M) associated with changes in viral copies. The results are based on the multivariate regression analysis using the sequence clusters (i.e., a categorical variable) inferred from the MDS components. (A) The phylogenetic tree estimated from a representative set of 996 genome sequences showing variant assignments and the locations of amino acid changes that increase (blue) or decrease (red) viral copies. (B) The temporal dynamics of the SNPs from February 2021 to March 2023. The transparency of the color corresponds to the mutation fraction in the daily sequence count: transparent color indicates low fractions, and opaque color indicates high fractions. The temporal dynamics of the SNPs, using MDS-inferred distance as a population control, are shown in **Fig. S9-11**.

To explore the temporal dynamics of these amino acid changes, we calculated the fraction of SNPs occurring in the sequences for each day, thereby accounting for the number of introductions to the population (**Fig. 4B**). We observed most SNPs with a positive impact on viral copies emerging in sequences sampled from February 2022, when BA.2 was first detected in Connecticut. These SNPs were consistently observed in almost every sequence thereafter. By contrast, we found that the other two amino acid changes (S:L452Q and S704L) that had a positive effect on viral copies were only in the samples from BA.2 infections and did not arise again in sublineages of BA.4 or BA.5. S:G252V was associated with higher viral copies; however, we found that the SNP only appeared in a few sequences associated with XBB infections. Notably, the N:R203M mutation was only associated with Delta infections. For the SNPs (ORF1ab:L5086I, S:N856K and L981F) that had a negative association with viral copies, we observed that they were present in samples associated with BA.1 infections and did not persist when BA.1 was replaced by BA.2.

## Discussion

We conducted a GWAS analysis on 9,902 high-quality SARS-CoV-2 genome sequences generated from two years of genomic surveillance in Connecticut, US to identify and evaluate SNPs that were associated with variations in viral copies during infections. Using a GWAS approach, we were able to identify and examine virus-related factors that were associated with the observed variations in viral copies independent of host factors. This was achieved by combining data from a large cohort of individuals infected with different VOCs and employing a regression model for viral copies that accounted for virus-level factors (i.e., specific SNPs and genetic background), adjusted for individual factors (i.e., age and vaccination status). We identified several SNPs corresponding to non-synonymous amino acid changes in the SARS-CoV-2 genome that were individually or jointly associated with the variations in viral copies. In particular, temporal patterns of the SNPs revealed that SNPs associated with increased viral copies were predominantly observed in Delta and Omicron BA.2/BA.4/BA.5/XBB infections, whereas those associated with decreased viral copies were mostly observed in infections with Omicron BA.1 variants.

Using a GWAS approach, we successfully identified a subset of variant-defining amino acid changes in Delta and Omicron variants (**Fig. S12**). Note that we did not detect any substitutions in the Alpha and Gamma variants (likely due to the low sample size for Gamma). We also identified SNPs that did not define any major variant category, including S:L452Q and S704L that were specifically associated with BA.2.12.1, a sublineage of BA.2 that briefly dominated during the pandemic (i.e., dominated mainly in the US between March and May 2022). This highlights the application of GWAS for identifying SNPs associated with important phenotypic effects without requiring a set of lineage-defining mutations to be defined a priori. Nevertheless, there are several reasons why we only detected a subset of the SNPs that defined different VOCs. Firstly, SNPs with small effect sizes may not be detected due to the stringent statistical significance thresholds applied in GWAS. Secondly, lineage-defining SNPs that are in low linkage disequilibrium with the causal mutations may not be detected [39], even if they may be functionally relevant. Our results showcase how GWAS can help to narrow the focus of SNPs associated with specific phenotypes, generating hypotheses for further investigation.

A key result from our analysis is that SNPs associated with viral copies did not exhibit the same temporal dynamics, even though they could have similar (either positive or negative) effects on viral copies, suggesting they may have independent effects on viral copies. Some amino acid changes, for example, ORF1ab:I4915T (positive effects), were only present in samples with BA.2 infections and disappeared when new Omicron variants emerged. Other SNPs (e.g., S:T19R), while also associated with higher viral copies, were observed and persisted in all BA.2/BA.4/BA.5/XBB infections. The distinct temporal pattern of SNPs, dependent on VOCs, may help explain the different fitness levels (e.g., intrinsic transmissibility or immune escape) of each variant [40,41]. Notably, we found three SNPs, ORF1ab:L5086I, S:N856K and L981F, were associated with decreased viral copies in BA.1 infections. The negative impact on viral copies should be interpreted with caution. Although the possibility that these SNPs have a direct impact on reducing viral copies cannot be ruled out, it is also likely that the estimated negative effects are due to a synthetic association with other SNPs. Further study may be required to disentangle the direct effects of these SNPs from the confounding influences of other genetic variations and to confirm their functional impact on viral copies.

In this study, we employed a series of single SNP regression models to identify the underlying SNPs associated with the changes in viral copies without accounting for potential interactions between SNPs. We noted that several synonymous SNPs located in the ORF1ab gene were identified to have an impact on viral copies. The synonymous SNPs were likely linked to non-synonymous SNPs that were under positive selection. In such cases, the synonymous SNPs can be carried along with the non-synonymous SNPs, resulting in their significance in the GWAS analysis, as shown in the subsequent epistasis test (**Fig. S3**). Nevertheless, the method provided an initial set of SNPs that are worth further exploration, pinpointing important mutations associated with viral copies and providing valuable insights into the overall genetic landscape of the viral population. The method, thus, represented an important first step towards understanding detailed epistatic effects among these mutations on viral copies. A paired or higher-order SNP regression study could be conducted as a subsequent step to test potential interactions or joint effects among different SNPs.

There are limitations to our study. First, we assumed that the distribution of times between infection and sample collection was similar through time and across variants as these data were not available. Given our samples were taken frequently over a 2-year period, we do not anticipate that this assumption will qualitatively impact our results. Second, our study primarily focuses on the genetic variants in VOCs, neglecting other factors such as host immune responses or environmental influences, partially captured by the host-associated covariates, including age and vaccination status in this study, that may also contribute to the changes in viral copies. Further study will be needed to address the impact of these factors on viral copies, for example, genome-to-genome analysis to reveal the impact of host-viral genetic interactions in SARS-CoV-2 infections [20,56]. Third, our data were obtained from a specific geographic region, whose population diversity may not necessarily be similar to other settings; therefore, extrapolating these findings to a broader population may require caution. Additionally, focusing solely on consensus genomic changes in the analysis could overlook the genetic diversity within the sample, which may also influence variations in viral load. Despite these constraints, our study highlights the importance of sustained genomic surveillance and the need for comprehensive analyses to understand the nuanced impact of specific genetic variations on viral copies at the within-host level, and its implications for viral transmissibility and immune escape at the population level. Further work and collaborative efforts are essential to elucidate the complex interplay between viral genetics, host factors, and the dynamics of transmission associated with emerging variants. Such studies could inform predictive early warning public health systems regarding the emergence of potentially highly transmissible viral strains based on their constellation of mutations.

Recently, Duesterwald et al. [12] used genome sequence data and a machine-learning approach to predict cycle threshold (Ct) values of SARS-CoV-2 infections based on the *k*-mers. Similar to our findings, they suggested that S:L452 and P681 were hallmarks of VOCs, implying impacts on the observed Ct values in clinical samples. Although the machine-learning approach may capture broader patterns and interactions within the genome on Ct values, they lack interpretability compared to regression models. For example, regression-based models could offer insights into the direct association between specific genetic variants and viral copies. In addition, regression-based models may perform well even with limited sample sizes [19], provided that the assumptions of the model are met and the predictors are informative, whereas using machine-learning methods with small sample sizes can be challenging. However, the viral GWAS method may not be appropriate in situations where there is insufficient genetic diversity in the viral population under study, as this can limit the power to detect meaningful associations between mutations and viral traits. Additionally, it may not be suitable when the phenotypic traits of interest are not well-defined or accurately measured.

With the availability of high-quality whole-genome sequences for SARS-CoV-2, we demonstrated that GWAS analysis of the viral genome can identify SNPs that associate with positive or negative impacts on viral copies in VOCs, revealing important biological insights and enhancing our understanding of within-host viral dynamics. We argue that the application of GWAS analyses to study viral genomes provides a particularly tractable tool to identify potential SNPs of interest for further evaluation across different viral pathogens. It is particularly useful to understand the genetic basis of viral virulence, transmission, resistance to antiviral treatments, and host-virus interactions for several reasons. First, the small genome size of viruses and high evolutionary rates make it easier to perform comprehensive genome-wide scans for SNPs and to experimentally test the impacts of SNPs on specific traits. Second, significant phenotypic variations (e.g., viral loads and antibody responses) are often observed in viral infections, despite limited changes in the viral genome. GWAS can help to identify SNPs that correlate with these phenotypic variations, providing insights into the genetic basis of these traits. Third, the increasing accessibility to sequence viral genomes makes it possible to perform GWAS on rich datasets, enabling in-depth analysis of the temporal dynamics of viral evolution. Together, the applicability of GWAS analyses to study viral genomes can provide a new approach for exploring the intricate interplay between genetic mutations and phenotypes, informing strategies for managing and mitigating the impact of emerging viral variants, and contributing to the development of potential therapeutic interventions.

## Materials & Methods

### Ethics

The Institutional Review Board from the Yale University Human Research Protection Program determined that the RT-qPCR testing and sequencing of de-identified remnant COVID-19 clinical samples obtained from clinical partners conducted in this study is not research involving human subjects (IRB Protocol ID: 2000028599).

### Clinical sample collection and measurement of viral copies by RT-qPCR

SARS-CoV-2 positive samples (nasal swabs in viral transport media) were collected through the Yale New Haven Hospital (YNHH) System as a part of routine inpatient and outpatient testing and sent to the Yale SARS-CoV-2 Genomic Surveillance Initiative. Using the MagMAX viral/pathogen nucleic acid isolation kit, nucleic acid was extracted from 300μl of each clinical sample and eluted into 75μl of elution buffer. Extracted nucleic acid was then used as template for a “research use only” (RUO) RT-qPCR assay [23] to test for presence of SARS-CoV-2 RNA. Ct values from the nucleocapsid target (CDC-N1 primer-probe set [57]) were used to derive viral copy numbers using a previously determined standard curve for this primer set [58]. A positive RNA control with defined viral copy number (1000/μl) was used to standardize results across individual runs.

### Whole genome sequencing

Libraries were prepared for sequencing using the Illumina COVIDSeq Test (RUO version) and quantified using the Qubit High Sensitivity dsDNA kit. Negative controls were included for RNA extraction, cDNA synthesis, and amplicon generation. Prepared libraries were sequenced at the Yale Center for Genomic Analysis on the Illumina NovaSeq with a 2x150 approach and at least 1 million reads per sample.

Reads were then aligned to the Wuhan-Hu-1 reference genome (GenBank MN908937.3) using BWA-MEM v.0.7.15 [59]. Adaptor sequences were then trimmed, primer sequences masked, and consensus genomes called (simple majority >60% frequency) using iVar v1.3.133 [60] and SAMtools v1.11 [61]. When <20 reads were present at a site an ambiguous “N” was used, with negative controls consisting of ≥99% Ns. The Pangolin lineage assignment tool [62] was used for assigning viral lineages.

### Clinical metadata

We obtained patient metadata and vaccination records from the YNHH system and the Center for Outcomes Research and Evaluation (CORE) and matched these records to sequencing data through unique sample identifiers. Duplicate patient records or those with missing or inconsistent metadata and vaccination date were removed from the GWAS analysis. We also removed patient records with persistent infections (>28 days since first positive test).

We determined vaccination status at time of infection by comparing the sample collection date to the patient’s vaccination record dates. We categorized vaccine statuses based on the number of vaccine doses received at least 14 days before the collection date. Patient vaccination statuses at the time of infection were categorized as follows: non-vaccine, one-dose vaccine, two-dose vaccine, two-dose vaccine with one booster, or two-dose vaccine with two boosters. We calculated the age of each patient as the difference between the date of birth and the sampling date.

### Single nucleotide polymorphisms

To identify single nucleotide polymorphisms (SNPs), we first aligned the 9902 genome sequences using *nextalign* (v3.2.1) [63] with the reference genome of the Wuhan-Hu-1 genome (GenBank accession: MN908937.3). Then, SNPs were identified using *snp-sites* (v2.4.1) [64], with the reference genome of the Wuhan-Hu-1 genome (GenBank accession: MN908937.3). We also normalized the SNPs in the generated VCF file, such that multiallelic SNPs were separated into different rows. Normalizing the SNPs ensured that each SNP was one-hot encoded and analyzed separately. Note that we did not include ambiguous SNPs, deletions and insertions in our GWAS analysis. We used *vcf-annotator* (v0.7) to annotate SNPs to corresponding amino acid changes.

### Multidimensional scaling and population control

To reveal the underlying structure of the 9902 genome sequences. We first used *snp-dists* (v0.7.0) [65] to convert the aligned sequences (a FASTA alignment) to a SNP distance matrix. We then applied a multidimensional scaling (MDS) method [26] to transform the SNP distance matrix into a geometric configuration while preserving the original pairwise relationships. The scaling was conducted using *cmdscale* function in an R package *stats* (v3.6.2). We set the maximal dimensional parameter *k* = 2.

To measure the goodness of the transformation, we calculated the distance between the original genome sequencing data and compared it with the new distances determined by MDS. This involved arranging the two matrices of distances into two columns and computing the correlation coefficient (i.e., *r*) between them. Finally, we used *r*^&^ to measure the proportion of variance in the original distance matrix explained by the new computed distance matrix.

To determine the clusters (i.e., categorical variables) from MDS, we applied the k-means clustering method using the kmeans function implemented in R statistical software (v4.0.2). We set the number of centroids *k* = 4.

### Testing for associations between viral copies and SNPs

In this work, we conducted a series of single SNP regression analyses to test for associations between viral copies and SNPs, adjusted for host ages, vaccination status and viral population. The linear regression model is written as follows:

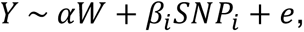

where *Y* is a vector of normalized log10-transformed viral copies, *W* is a matrix of covariates, including age (a categorical variable with four age groups of “<18”, “18-49”, “50-69”, and “>70” years old), vaccination status (a categorical variable with vaccination statuses of “0 doses”, “1 dose”, “2 doses”, “2 doses and 1 booster”, “2 doses and 2 boosters”), a population control variable for different viral variants (a categorical variable with cluster numbers of “1”, “2”, “3” and “4”, see **Fig. S1** for detailed clusters), and an intercept, and *α* is a vector that corresponds to coefficients of the covariates. In particular, *SNP_i_* is a vector of genotype values for all samples at each SNP, *i*. It is a binary variable: 0 represents the SNP is not present in the genome sequence, whereas 1 represents its presence. *β_i_* is the effective size of each identified SNP, *i*. We also conducted a sensitivity analysis including two terms *ξ*_1_*d*_1_, *ξ*_2_*d*_2_ as covariates in the model for population control. The vectors *d*_1_, *d*_2_ represent the two dimensions computed by MDS, and *ξ*_1_, *ξ*_2_ are the coefficients of the dimension covariates. The random effect of residual errors is presented here by *e*, which is assumed to follow a normal distribution with a mean of 0 and a standard deviation of *σ_e_*.

### Marginal epistasis test

We applied a marginal epistasis test method to explore the interactions between SNPs on viral copies, using an R package mvMAPIT (v.2.0.3) [28–30]. This method maps SNPs with non-zero marginal epistatic effects—the combined pairwise interaction effects between a given SNP and all other SNPs—identifying candidate variants involved in epistasis without needing to identify the exact partners with which the variants interact.

### Phylogenetic tree construction and comparison to variant-defining substitutions

We employed *iq-tree* (v2.2.2.6) [66] of a representative set using 996 of our 9902 genome sequences for tree construction, using Wuhan-Hu-1 (GenBank MN908937.3) as the reference genome. We specified the HKY substitution model and set the number of bootstrap replicates to 1,000. To visualize the phylogenetic tree, we used the ggtree (v1.4.11) implemented in the R statistical software (v4.0.2). The variant-defining amino acid changes were defined as those mutations with >75% prevalence in at least one lineage, as estimated on outbreak.info website [38]. Note that we did not include deletions in variant-defining substitutions.

## Data Availability

Data and code used in this study are publicly available on Github: https://github.com/grubaughlab/2024_paper_GWAS. All genome sequences used for the GWAS analysis and a subset of the associated metadata (accession number, virus name, collection date, originating lab and submitting lab, and the list of authors) in this dataset are published in GISAIDs EpiCoV database: https://doi.org/10.55876/gis8.240219fh. The de-identified and coded clinical metadata associated with the sequenced samples are available upon request with IRB approval.

## Data and code availability

We used the R statistical software (v4.0.2) for all statistical analyses and visualization. Data and code used in this study are publicly available on Github: https://github.com/grubaughlab/2024_paper_GWAS. All genome sequences used for the GWAS analysis and a subset of the associated metadata (accession number, virus name, collection date, originating lab and submitting lab, and the list of authors) in this dataset are published in GISAID’s EpiCoV database: https://doi.org/10.55876/gis8.240219fh. The de-identified and coded clinical metadata associated with the sequenced samples are available upon request with IRB approval.

## Acknowledgements

We would like to thank Verity Hill, Seth Redmond, Jiye Kwon, Rafael Lopes, Sophie Taylor, and Philip Jack for their helpful conversations and feedback on this work. This project is supported by the Centers for Disease Control and Prevention (CDC) Broad Agency Announcement Contract 75D30122C14697 (NDG). This work does not necessarily represent the views of the CDC.

## Yale SARS-CoV-2 Genomic Surveillance Initiative Authors

Tara Alpert, Kaya Bilguvar, Kendall Billig, Mallery Breban, Anderson Brito, Christopher Castaldi, Rebecca Earnest, Bony De Kumar, Joseph Fauver, Chaney Kalinich, Tobias Koch, Marie Landry, Shrikant Mane, Isabel Ott, David Peaper, Mary Petrone, Kien Pham, Jessica Rothman, Irina Tikhonova, Chantal Vogels, Anne Watkins

## Supplementary Figures & Tables

**Supplemental Figure 1. Results of multidimensional scaling.** The population structure of the 9902 genome sequences using a multidimensional scaling (MDS) method. Clusters are defined using a *k*-means clustering method, as demonstrated on the bottom right corner.

**Supplemental Figure 2. Q-Q plots of GWAS p-values.** Q-Q plots (quantile-quantile plots) showing the p-values from GWAS analysis using **(A)** two MDS-computed components, or **(B)** MDS-inferred four clusters as covariates in the regression model.

**Supplemental Figure 3. Marginal epistasis tests identify single nucleotide polymorphisms (SNPs) that have epistatic interactions with others and are associated with the changes in viral copies.** (A) Marginal epistasis test results of the SNPs (annotated as amino acid changes) that have marginal epistatic effects on viral copies. The dashed line indicates the permuted threshold for genome-wide significance *p* = 0.05/171 = 2.74 × 10^-4^. Significant mutations are shown with solid colors. (B) The *p*-values and (C) the effect size of pairwise interaction tests among the significant mutations.

**Supplemental Figure 4. GWAS analysis identifies several single nucleotide polymorphisms (SNPs) that are associated with the changes in viral copies.** (A) Genome-wide association results of the impact of identified SNPs on viral copies during SARS-CoV-2 infection. The dashed line indicates the permuted threshold for genome-wide significance *p* = 4.67 × 10^-6^. Significant SNPs are shown with solid colors. (B) SNPs (with *p* < 1 × 10^-10^) that have positive (blue) or negative (red) effects on viral copies. (C) The corresponding synonymous (triangles) and non-synonymous (circles) amino acid changes that associate with increased or decreased viral copies. Data is shown as means with 95% confidence intervals. The estimated effective sizes and associated standard deviations are given in Table S1. A Q-Q plot showing the observed distribution of p-value and the expected distribution is given in **Fig. S2**.

**Supplemental Figure 5. GWAS analysis using only Cluster 1 data (shown in Fig. S1).** (**A**) Genome-wide association results of the impact of identified SNPs on viral copies during SARS-CoV-2 infection. The dashed line indicates the permuted threshold for genome-wide significance *p* = 4.03 × 10^-10^ (0.05/1242 SNPs). Significant SNPs are shown with solid colors. (**B**) SNPs (with *p* < 1 × 10 ^−10^) that have positive (blue) or negative (red) effects on viral copies. (**C**) The corresponding synonymous (triangles) amino acid changes that associate with increased or decreased viral copies. Data shown as means with 95% confidence intervals.

**Supplemental Figure 6. GWAS analysis using Cluster 2 data (shown in Fig. S1).** (**A**) Genome-wide association results of the impact of identified SNPs on viral copies during SARS-CoV-2 infection. The dashed line indicates the permuted threshold for genome-wide significance *p* = 3.68 × 10^-5^ (0.05/1357 SNPs). Significant SNPs are shown with solid colors. (**B**) SNPs (with *p* < 1 × 10^-10^) that have positive (blue) or negative (red) effects on viral copies. (**C**) The corresponding synonymous (triangles) amino acid changes that associate with increased or decreased viral copies. Data shown as means with 95% confidence intervals.

**Supplemental Figure 7. GWAS analysis using Cluster 3 data (shown in Fig. S1).** (**A**) Genome-wide association results of the impact of identified SNPs on viral copies during SARS-CoV-2 infection. The dashed line indicates the permuted threshold for genome-wide significance *p* = 2.80 × 10^-5^ (0.05/1784 SNPs). Significant SNPs are shown with solid colors. (**B**) SNPs (with *p* < 1 × 10^-10^) that have positive (blue) or negative (red) effects on viral copies. (**C**) The corresponding non-synonymous (circles) amino acid changes that associate with increased or decreased viral copies. Data shown as means with 95% confidence intervals.

**Supplemental Figure 8. GWAS analysis using Cluster 4 data (shown in Fig. S1).** (**A**) Genome-wide association results of the impact of identified SNPs on viral copies during SARS-CoV-2 infection. The dashed line indicates the permuted threshold for genome-wide significance *p* = 7.91 × 10^-6^ (0.05/6314 SNPs). Significant SNPs are shown with solid colors. (**B**) SNPs (with *p* < 1 × 10^-10^) that have positive (blue) or negative (red) effects on viral copies. (**C**) The corresponding synonymous (triangles) and non-synonymous (circles) amino acid changes that associate with increased or decreased viral copies. Data shown as means with 95% confidence intervals.

**Supplemental Figure 9. The temporal dynamics of amino acid changes in the S gene associated with changes in viral copies.** The results are based on the multivariate regression analysis using the two MDS components as covariates. (A) The phylogenetic tree estimated from a representative set of 996 genome sequences showing variant assignments and the locations of amino acid changes that increase (blue) or decrease (red) viral copies. (B) The temporal dynamics of the SNPs from February 2021 to March 2023. The transparency of the color corresponds to the mutation fraction in the daily sequence count: transparent color indicates low fractions, and opaque color indicates high fractions.

**Supplemental Figure 10. The temporal dynamics of amino acid changes in the ORF1ab gene associated with changes in viral copies.** The results are based on the multivariate regression analysis using the two MDS components as covariates. (A) The phylogenetic tree estimated from a representative set of 996 genome sequences showing variant assignments and the locations of amino acid changes that increase (blue) or decrease (red) viral copies. (B) The temporal dynamics of the SNPs from February 2021 to March 2023. The transparency of the color corresponds to the mutation fraction in the daily sequence count: transparent color indicates low fractions, and opaque color indicates high fractions.

**Supplemental Figure 11. The temporal dynamics of amino acid changes in the ORF3a gene (S26L and T223I), M gene (D3G and I82T), ORF7b gene (T40I) and N gene (D63G and S413R) associated with changes in viral copies.** The results are based on the multivariate regression analysis using the two MDS components as covariates. (A) The phylogenetic tree estimated from a representative set of 996 genome sequences showing variant assignments and the locations of amino acid changes that increase (blue) or decrease (red) viral copies. (B) The temporal dynamics of the SNPs from February 2021 to March 2023. The transparency of the color corresponds to the mutation fraction in the daily sequence count: transparent color indicates low fractions, and opaque color indicates high fractions.

**Supplemental Figure 12. Comparison of key variant-defining amino acid changes with GWAS-identified substitutions.** The comparison of the key amino acid changes (dark purple) in each variant, with GWAS-identified SNPs that were associated with negative (red) or positive (blue) effects on viral copies in the (A) S gene and (B) ORF1ab gene. The results of GWAS analysis using the two dimensions computed by MDS as covariates are shown as “GWAS 1”, and the results of the analysis using the categorical clusters as covariates are shown as “GWAS 2”. The effective sizes of identified SNPs using different population control methods are given in Tables S1 and S2.

**Supplemental Table 1. The identified amino acid changes associated estimated effective sizes and standard deviations using the multivariate linear regression model with categorical clusters as covariates.**

**Supplemental Table 2. The identified amino acid changes associated estimated effective sizes and standard deviations using the multivariate linear regression model with MDS-computed dimensions as covariates.**

