## Supplemental Figures and Text for "Genome-wide association study between SARS-CoV-2 single nucleotide polymorphisms and virus copies during infections": Supporting Information.docx

### Supplementary Figures & Tables

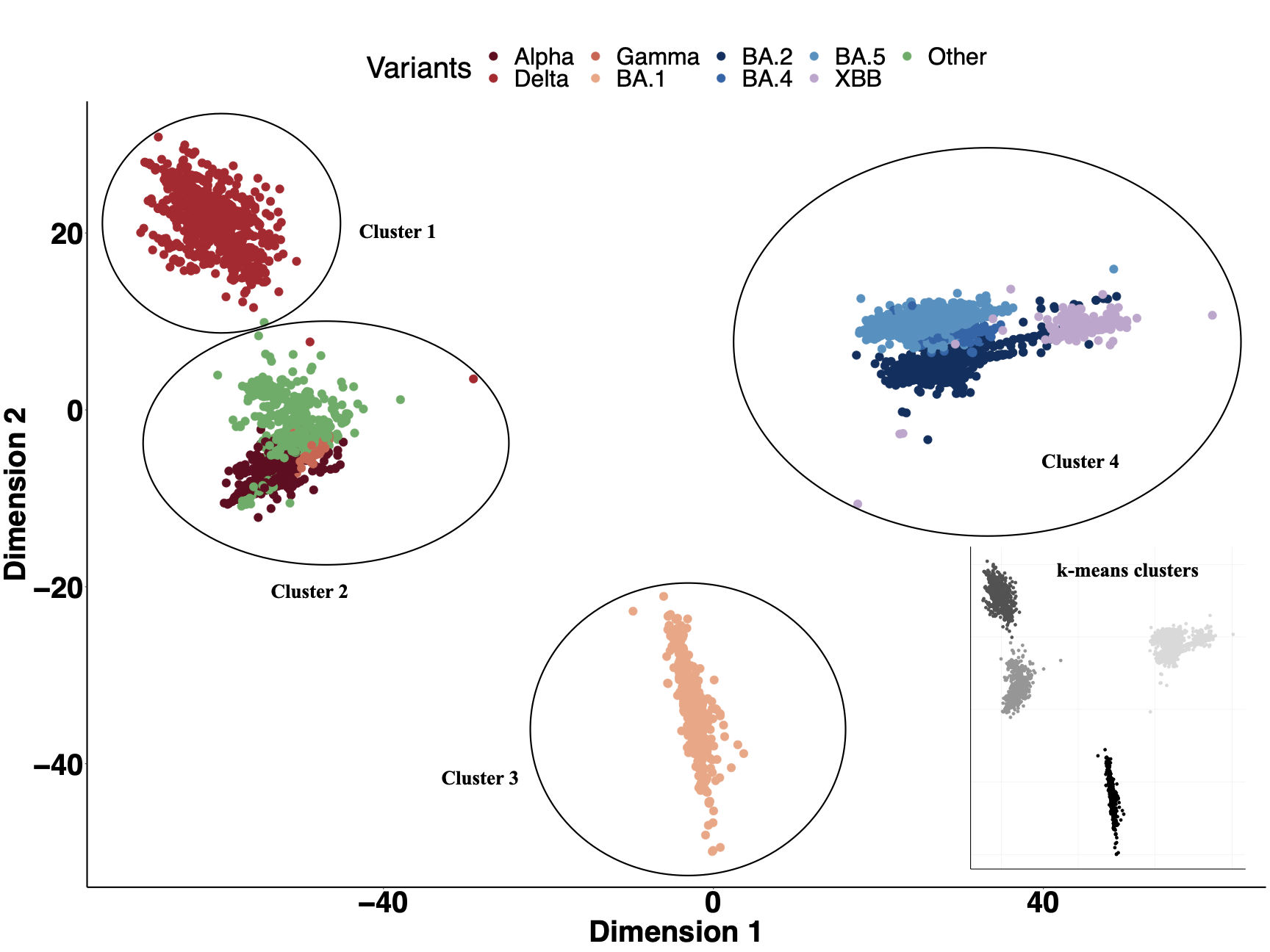

**Supplemental Figure 1. Results of multidimensional scaling.** The population structure of the 9902 genome sequences using a multidimensional scaling (MDS) method. Clusters are defined using a *k*-means clustering method, as demonstrated on the bottom right corner.

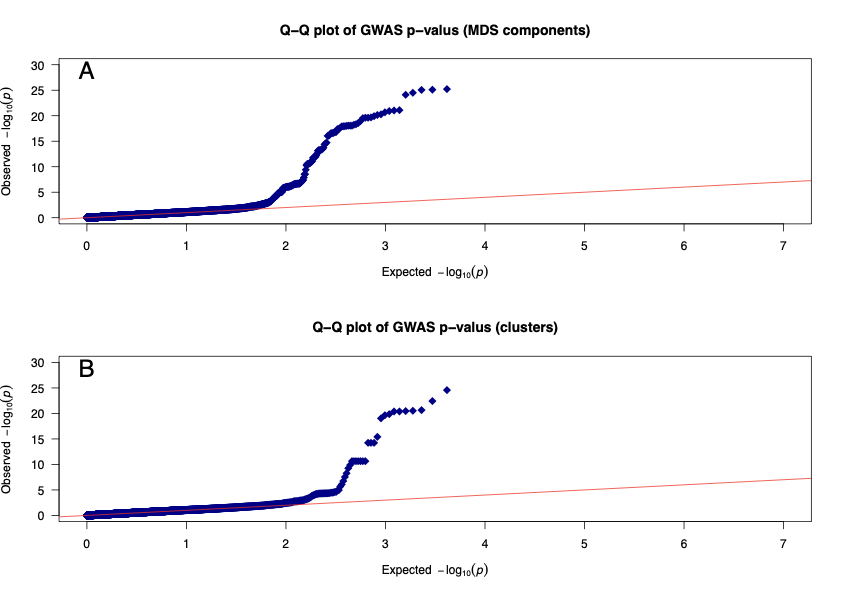

**Supplemental Figure 2. Q-Q plots of GWAS p-values.** Q-Q plots (quantile-quantile plots) showing the p-values from GWAS analysis using **(A)** two MDS-computed components, or **(B)** MDS-inferred four clusters as covariates in the regression model.

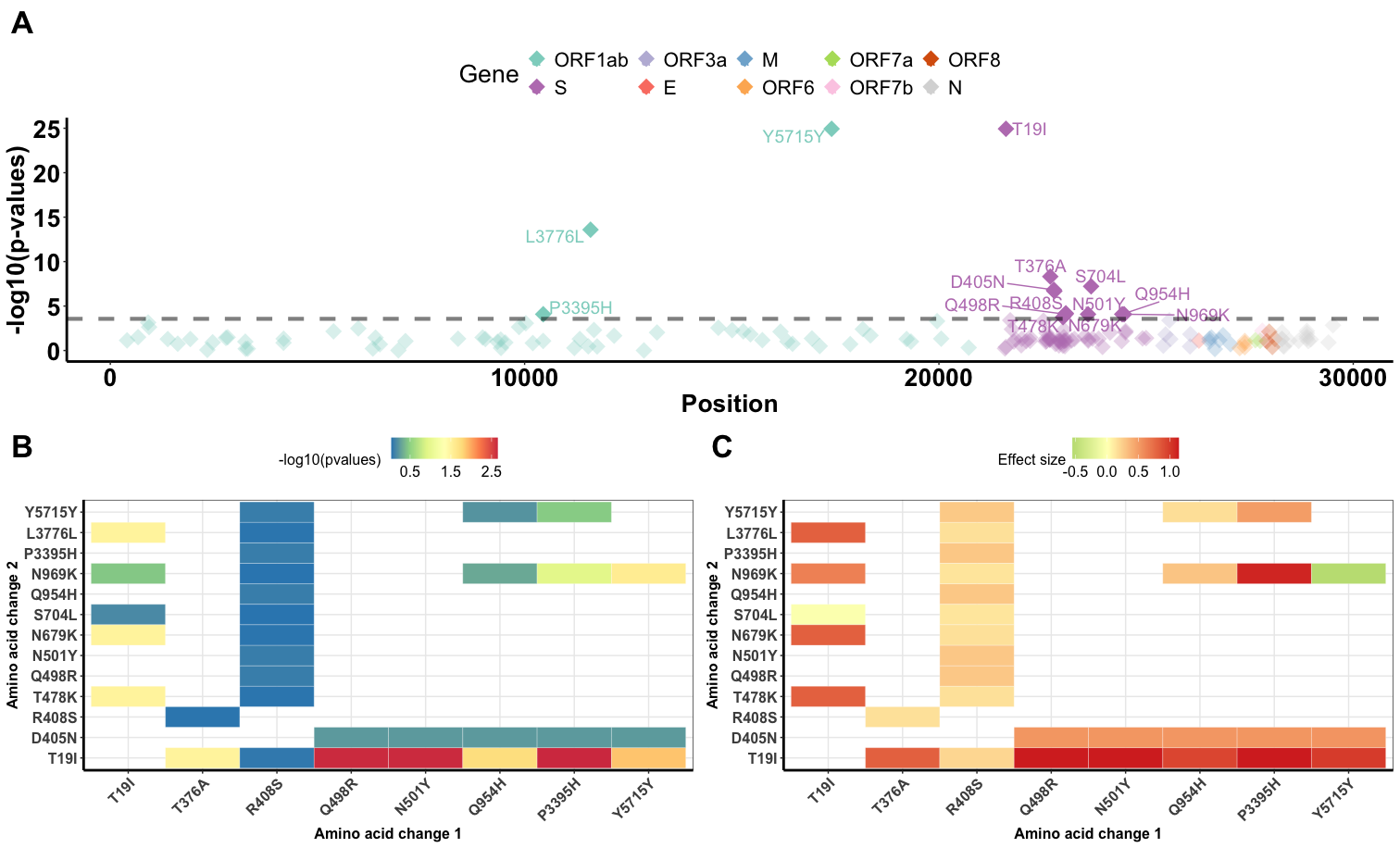

**Supplemental Figure 3. Marginal epistasis tests identify single nucleotide polymorphisms (SNPs) that have epistatic interactions with others and are associated with the changes in viral copies.** (A) Marginal epistasis test results of the SNPs (annotated as amino acid changes) that have marginal epistatic effects on viral copies. The dashed line indicates the permuted threshold for genome-wide significance $p$ = $0.05/171=2.74\times10^{-4}$. Significant mutations are shown with solid colors. (B) The $p$-values and (C) the effect size of pairwise interaction tests among the significant mutations.

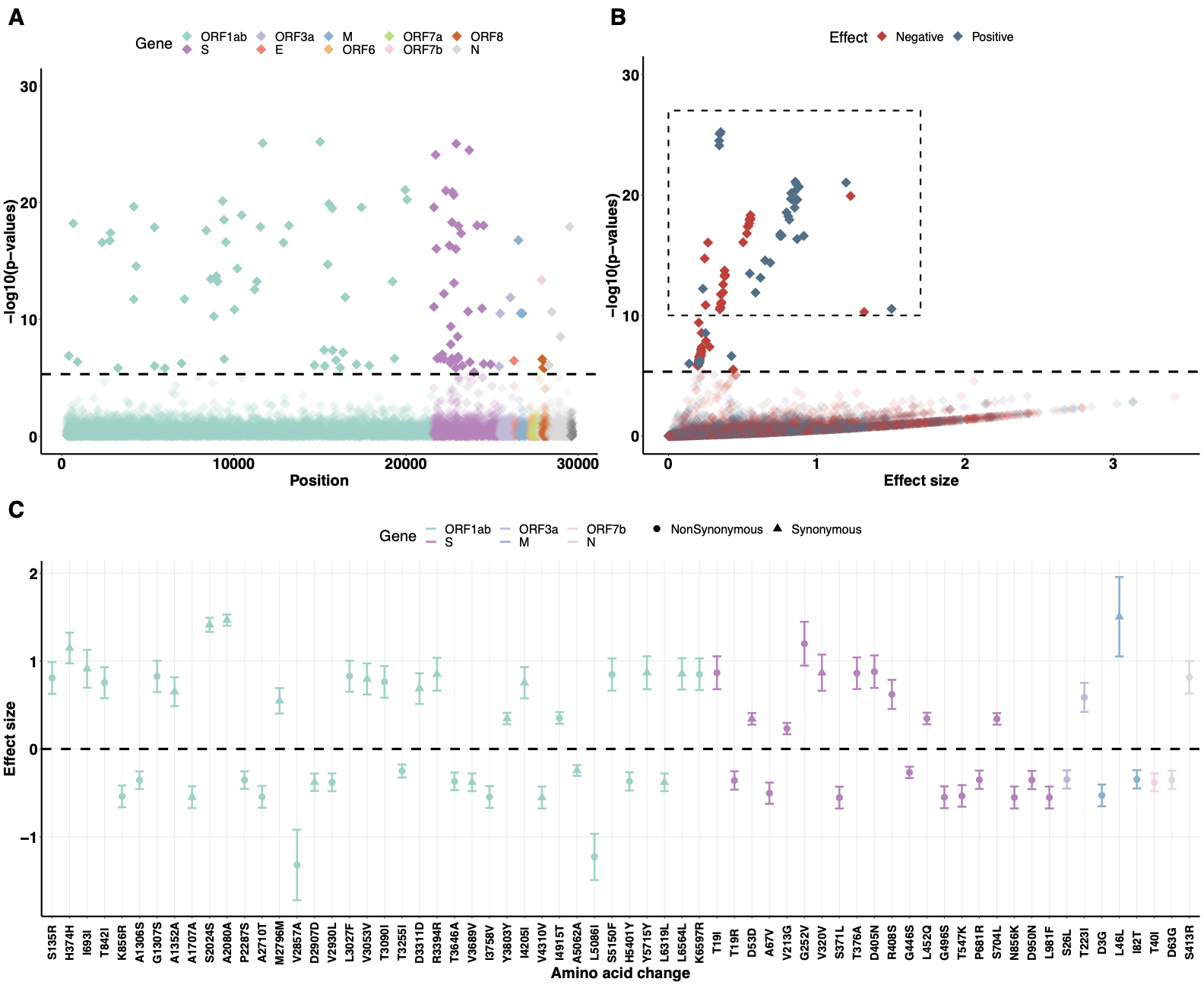

**Supplemental Figure 4. GWAS analysis identifies several single nucleotide polymorphisms (SNPs) that are associated with the changes in viral copies.** (A) Genome-wide association results of the impact of identified SNPs on viral copies during SARS-CoV-2 infection. The dashed line indicates the permuted threshold for genome-wide significance $p$ = $4.67\times10^{-6}$. Significant SNPs are shown with solid colors. (B) SNPs (with $p$ < $1\times10^{-10}$) that have positive (blue) or negative (red) effects on viral copies. (C) The corresponding synonymous (triangles) and non-synonymous (circles) amino acid changes that associate with increased or decreased viral copies. Data is shown as means with 95% confidence intervals. The estimated effective sizes and associated standard deviations are given in Table S1. A Q-Q plot showing the observed distribution of p-value and the expected distribution is given in **Fig. S2**.

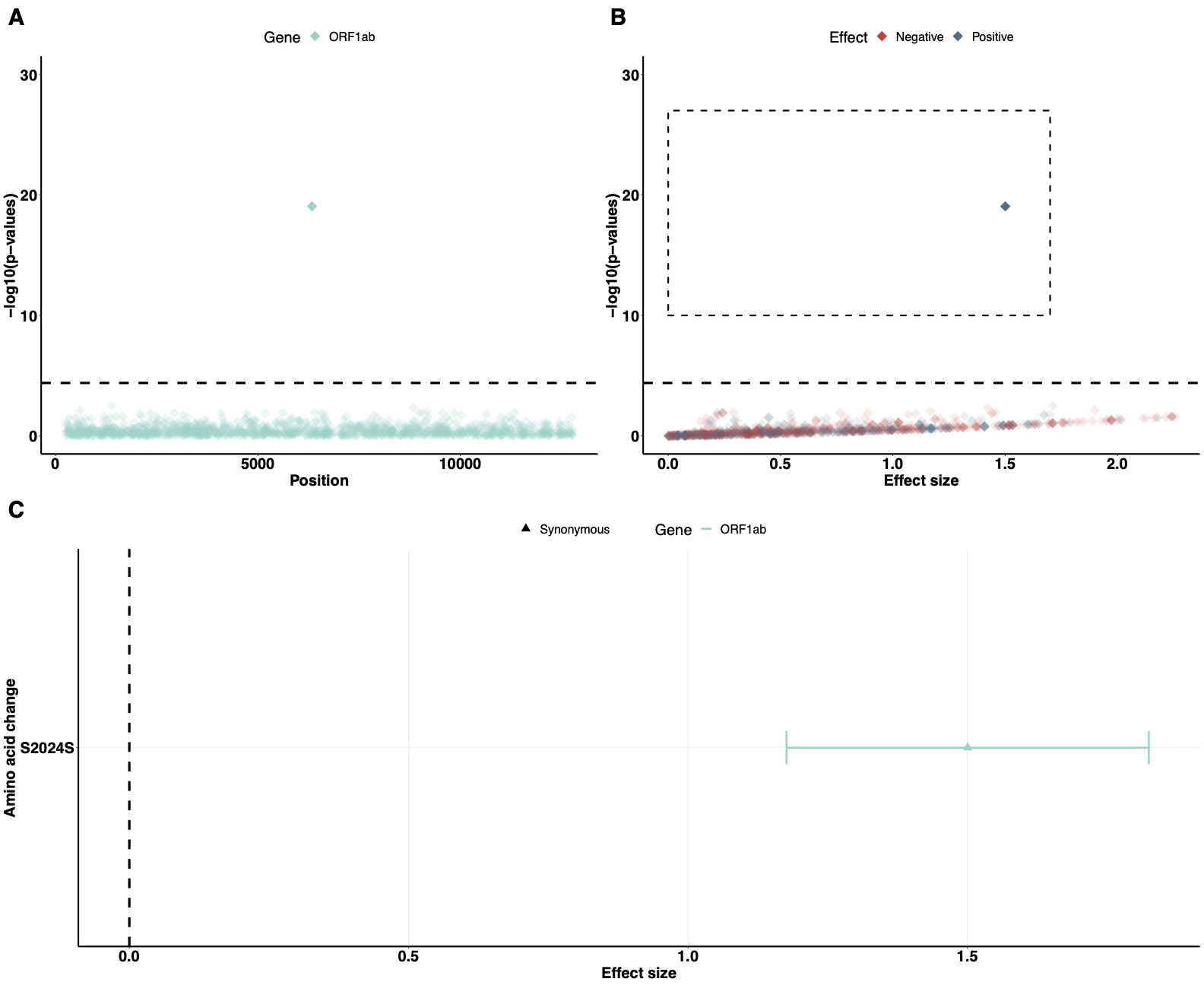

**Supplemental Figure 5. GWAS analysis using only Cluster 1 data (shown in Fig. S1).** (**A**) Genome-wide association results of the impact of identified SNPs on viral copies during SARS-CoV-2 infection. The dashed line indicates the permuted threshold for genome-wide significance $p$ = $4.03\times10^{-5}$ (0.05/1242 SNPs). Significant SNPs are shown with solid colors. (**B**) SNPs (with $p$ < $1\times10^{-10}$) that have positive (blue) or negative (red) effects on viral copies. (**C**) The corresponding synonymous (triangles) amino acid changes that associate with increased or decreased viral copies. Data shown as means with 95% confidence intervals.

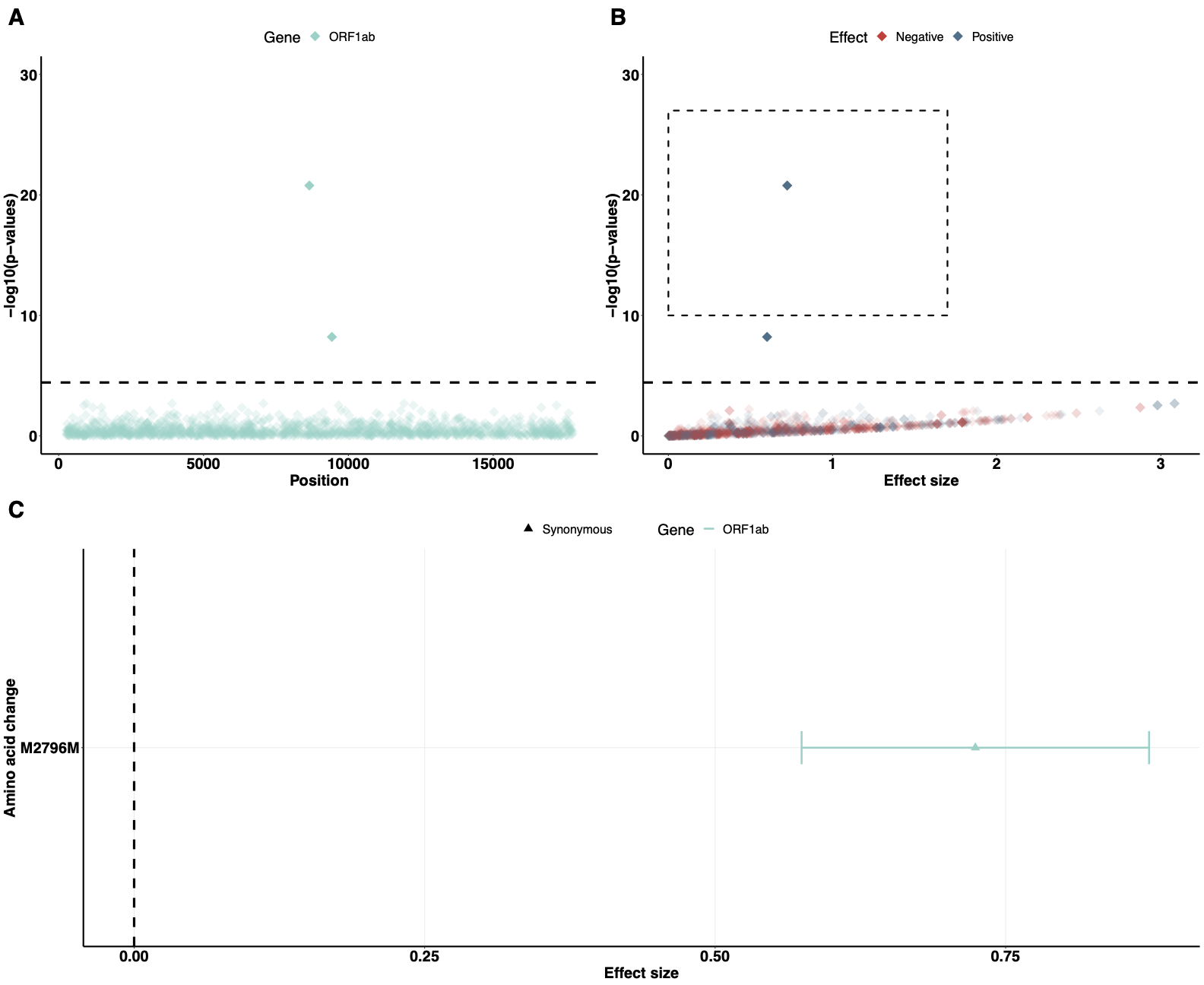

**Supplemental Figure 6. GWAS analysis using Cluster 2 data (shown in Fig. S1).** (**A**) Genome-wide association results of the impact of identified SNPs on viral copies during SARS-CoV-2 infection. The dashed line indicates the permuted threshold for genome-wide significance $p$ = $3.68\times10^{-5}$(0.05/1357 SNPs). Significant SNPs are shown with solid colors. (**B**) SNPs (with $p$ < $1\times10^{-10}$) that have positive (blue) or negative (red) effects on viral copies. (**C**) The corresponding synonymous (triangles) amino acid changes that associate with increased or decreased viral copies. Data shown as means with 95% confidence intervals.

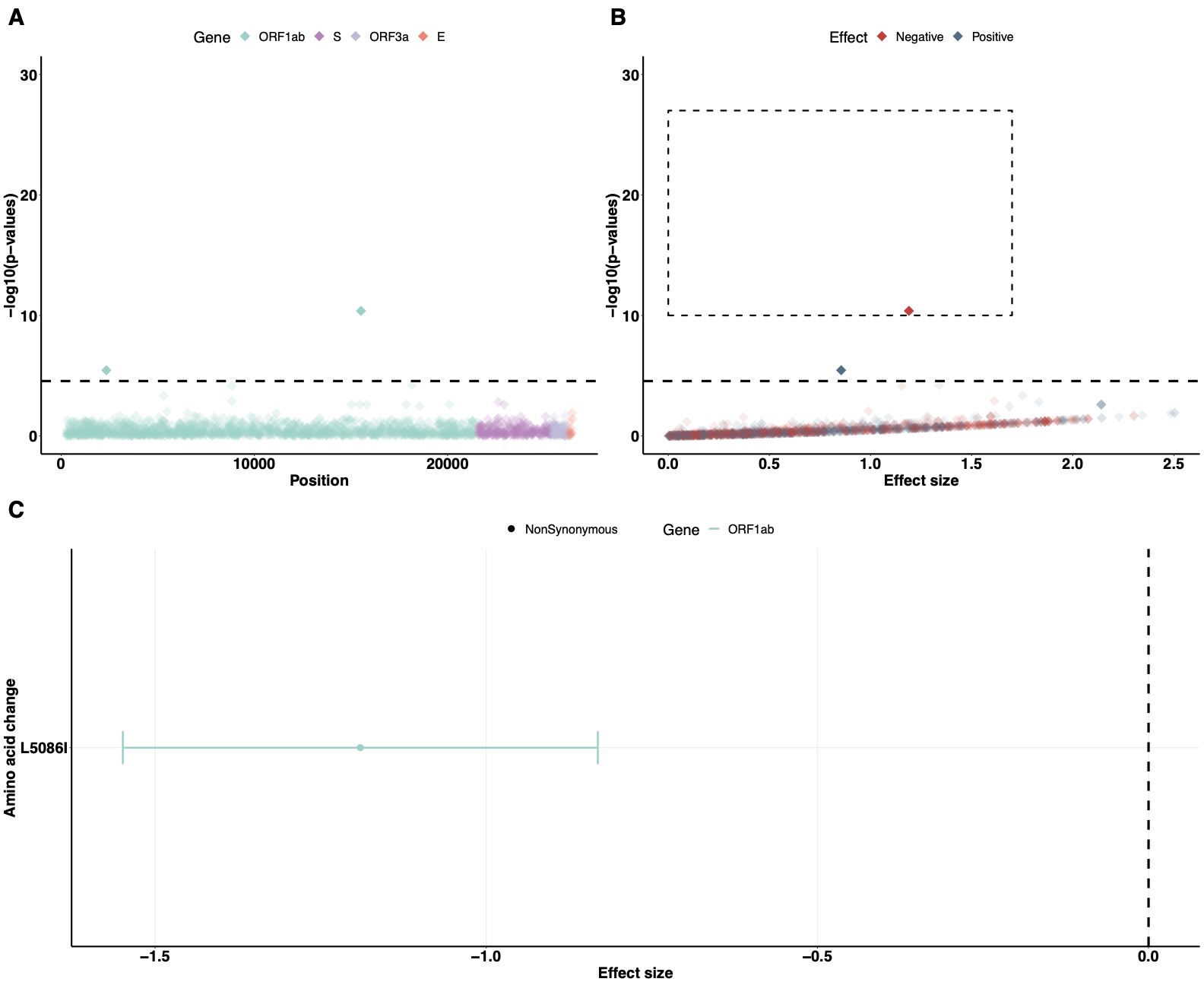

**Supplemental Figure 7. GWAS analysis using Cluster 3 data (shown in Fig. S1).** (**A**) Genome-wide association results of the impact of identified SNPs on viral copies during SARS-CoV-2 infection. The dashed line indicates the permuted threshold for genome-wide significance $p$ = $2.80\times10^{-5}$(0.05/1784 SNPs). Significant SNPs are shown with solid colors. (**B**) SNPs (with $p$ < $1\times10^{-10}$) that have positive (blue) or negative (red) effects on viral copies. (**C**) The corresponding non-synonymous (circles) amino acid changes that associate with increased or decreased viral copies. Data shown as means with 95% confidence intervals.

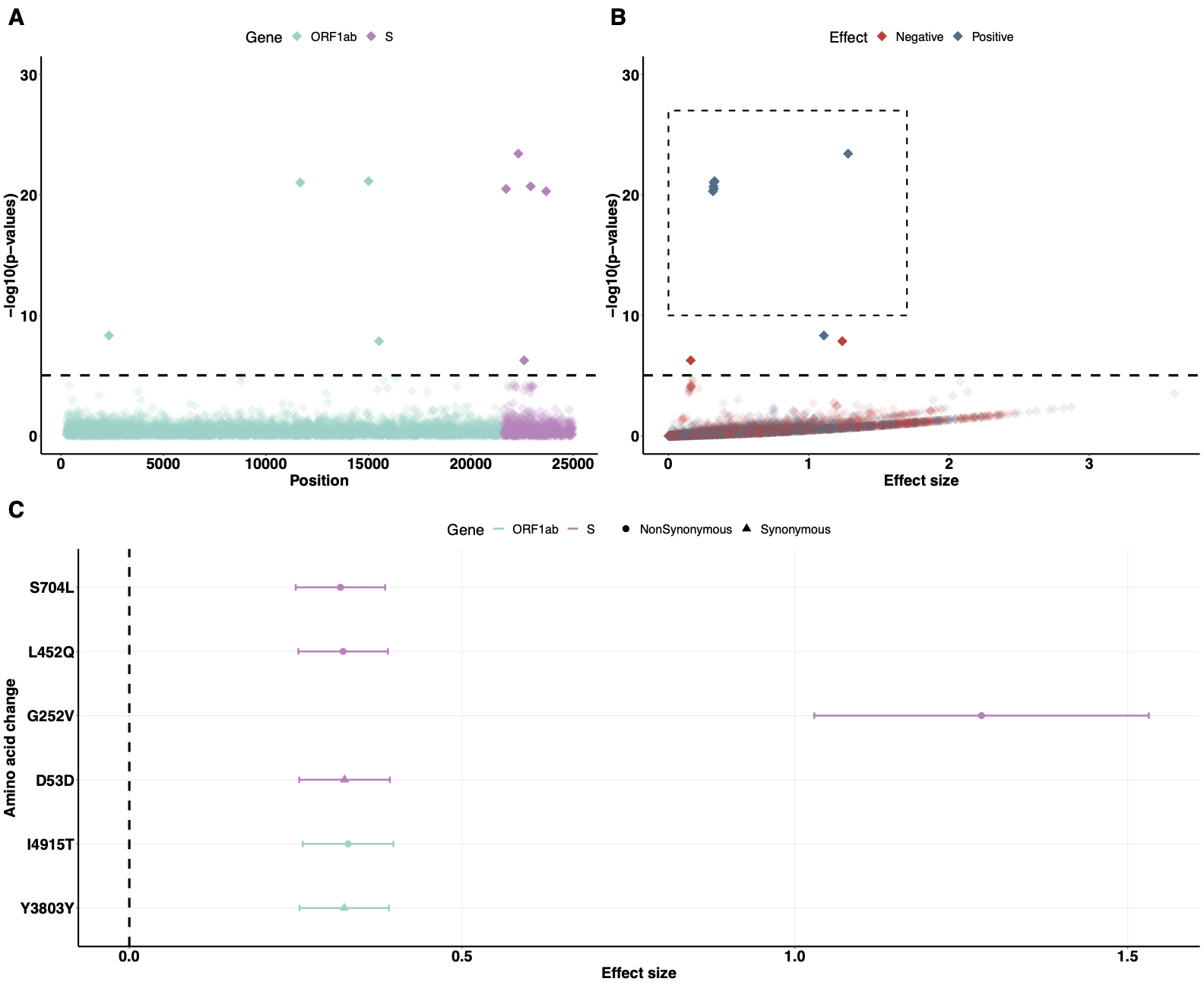

**Supplemental Figure 8. GWAS analysis using Cluster 4 data (shown in Fig. S1).** (**A**) Genome-wide association results of the impact of identified SNPs on viral copies during SARS-CoV-2 infection. The dashed line indicates the permuted threshold for genome-wide significance $p$ = $7.91\times10^{-6}$ (0.05/6314 SNPs). Significant SNPs are shown with solid colors. (**B**) SNPs (with $p$ < $1\times10^{-10}$) that have positive (blue) or negative (red) effects on viral copies. (**C**) The corresponding synonymous (triangles) and non-synonymous (circles) amino acid changes that associate with increased or decreased viral copies. Data shown as means with 95% confidence intervals.

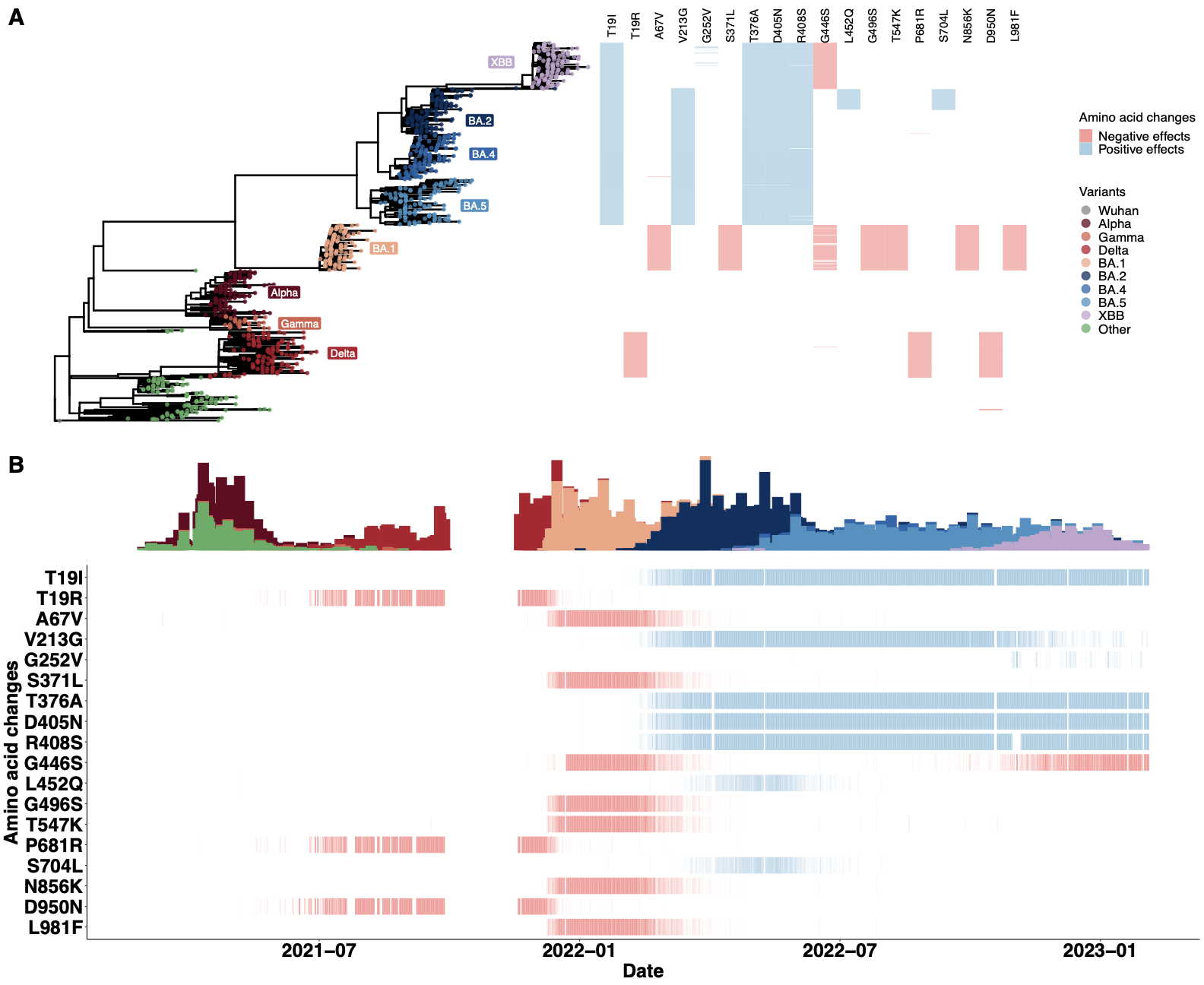

**Supplemental Figure 9. The temporal dynamics of amino acid changes in the S gene associated with changes in viral copies.** The results are based on the multivariate regression analysis using the two MDS components as covariates. (A) The phylogenetic tree estimated from a representative set of 996 genome sequences showing variant assignments and the locations of amino acid changes that increase (blue) or decrease (red) viral copies. (B) The temporal dynamics of the SNPs from February 2021 to March 2023. The transparency of the color corresponds to the mutation fraction in the daily sequence count: transparent color indicates low fractions, and opaque color indicates high fractions.

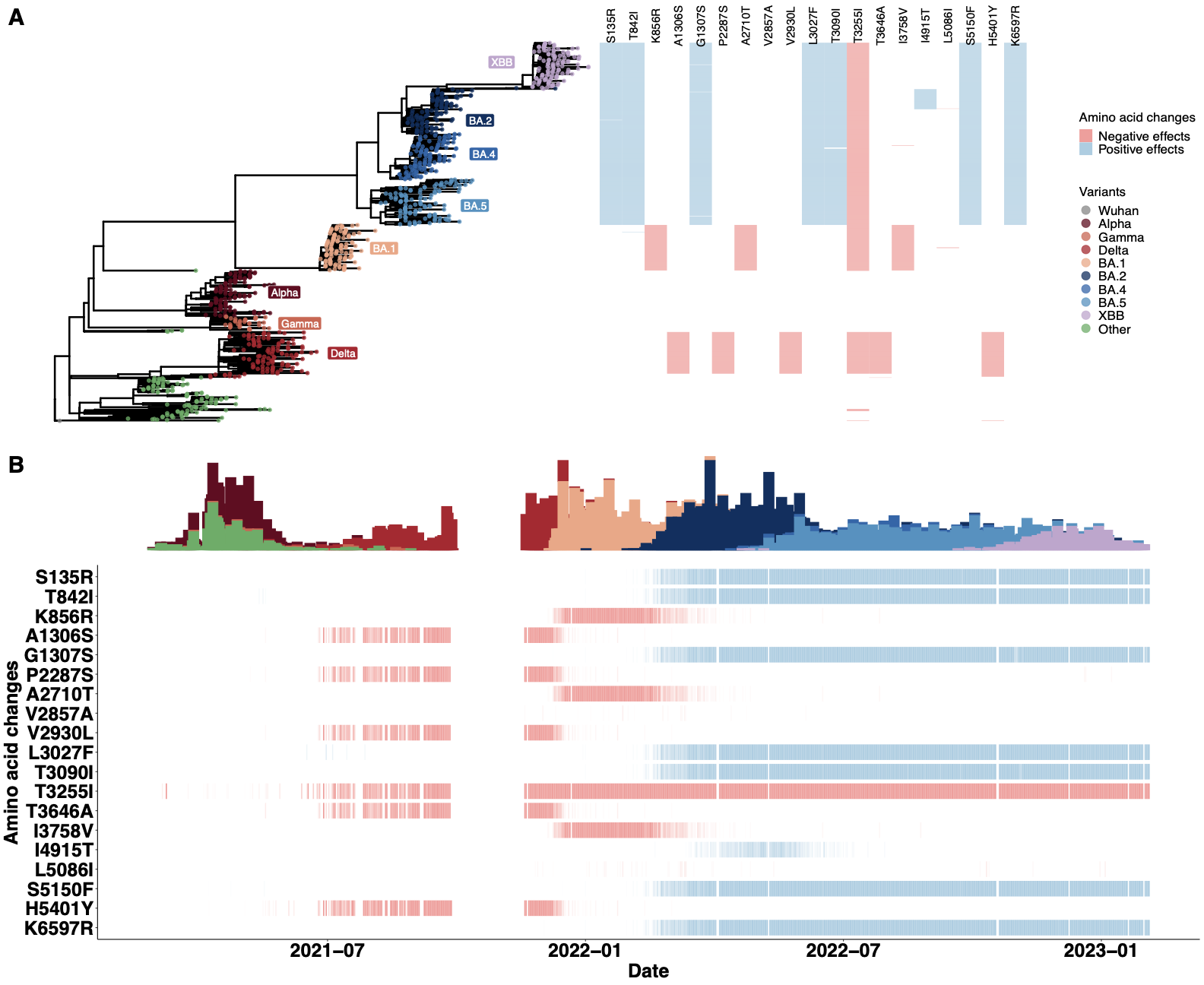
**Supplemental Figure 10. The temporal dynamics of amino acid changes in the ORF1ab gene associated with changes in viral copies.** The results are based on the multivariate regression analysis using the two MDS components as covariates. (A) The phylogenetic tree estimated from a representative set of 996 genome sequences showing variant assignments and the locations of amino acid changes that increase (blue) or decrease (red) viral copies. (B) The temporal dynamics of the SNPs from February 2021 to March 2023. The transparency of the color corresponds to the mutation fraction in the daily sequence count: transparent color indicates low fractions, and opaque color indicates high fractions.

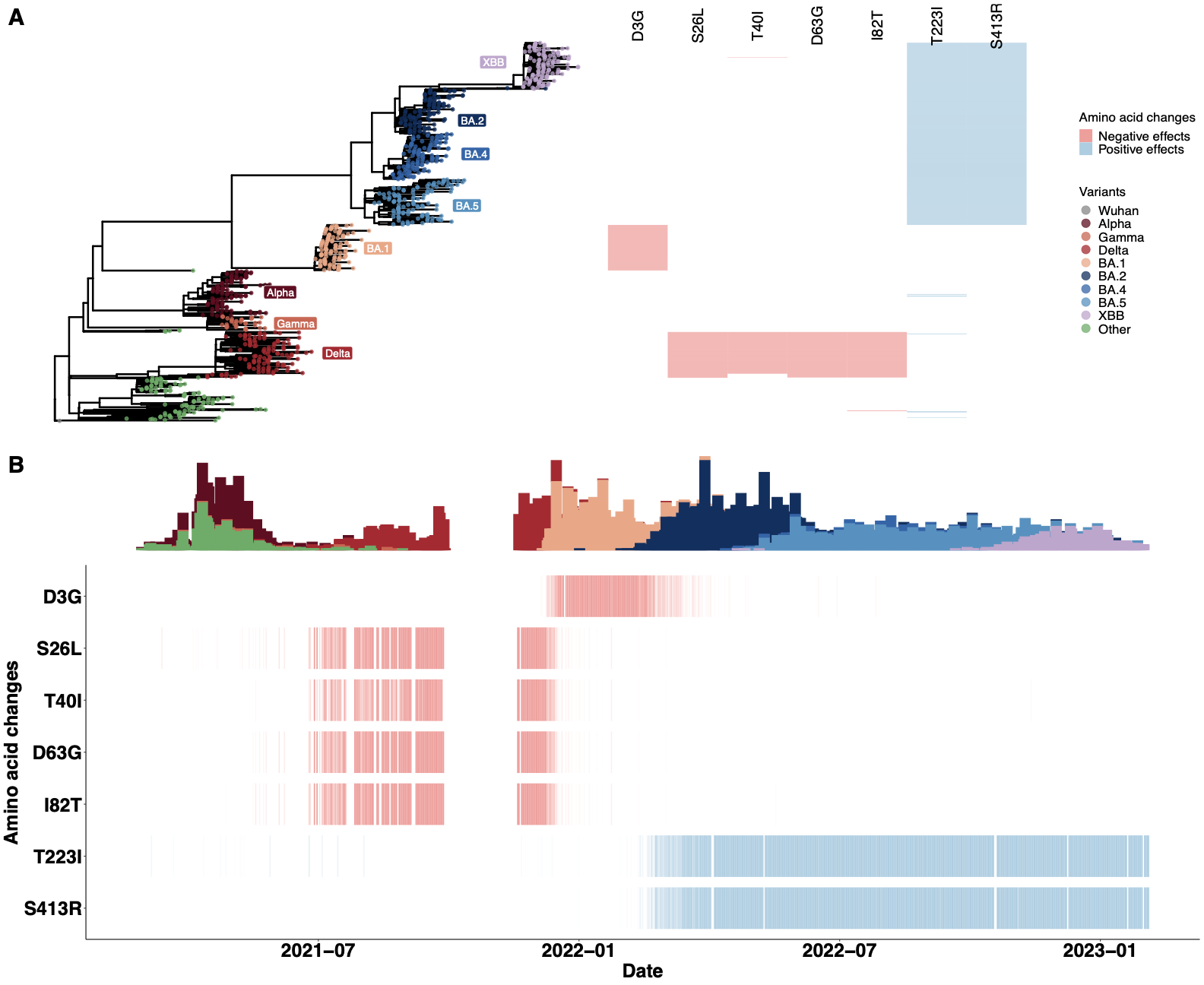

**Supplemental Figure 11. The temporal dynamics of amino acid changes in the ORF3a gene (S26L and T223I), M gene (D3G and I82T), ORF7b gene (T40I) and N gene (D63G and S413R) associated with changes in viral copies.** The results are based on the multivariate regression analysis using the two MDS components as covariates. (A) The phylogenetic tree estimated from a representative set of 996 genome sequences showing variant assignments and the locations of amino acid changes that increase (blue) or decrease (red) viral copies. (B) The temporal dynamics of the SNPs from February 2021 to March 2023. The transparency of the color corresponds to the mutation fraction in the daily sequence count: transparent color indicates low fractions, and opaque color indicates high fractions.

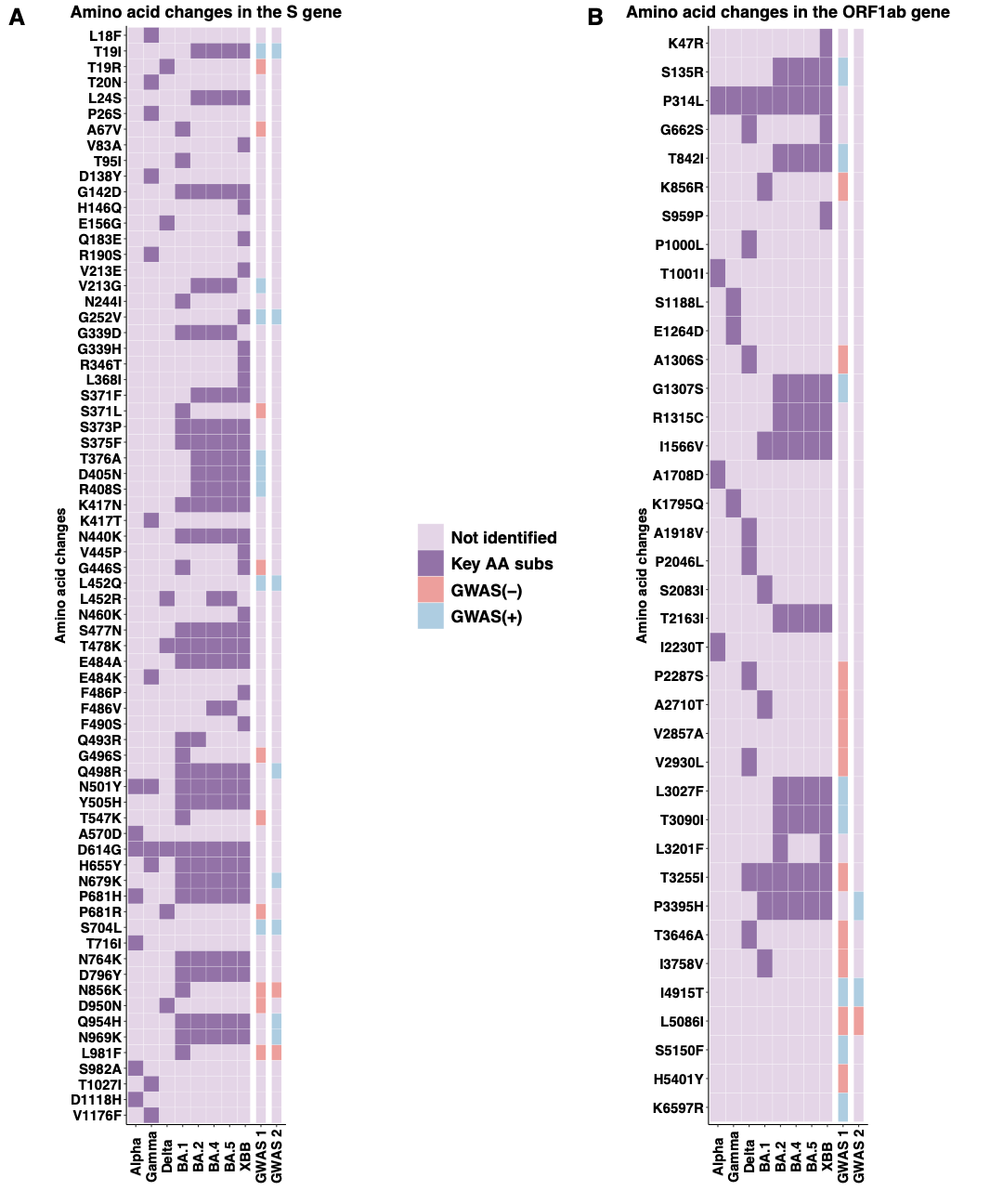

**Supplemental Figure 12. Comparison of key variant-defining amino acid changes with GWAS-identified substitutions.** The comparison of the key amino acid changes (dark purple) in each variant, with GWAS-identified SNPs that were associated with negative (red) or positive (blue) effects on viral copies in the (A) S gene and (B) ORF1ab gene. The results of GWAS analysis using the two dimensions computed by MDS as covariates are shown as “GWAS 1”, and the results of the analysis using the categorical clusters as covariates are shown as “GWAS 2”. The effective sizes of identified SNPs using different population control methods are given in Tables S1 and S2.

| AminoAcidChange | coefficients | Standard deviation |
| --- | --- | --- |
| T19I | 0.2123842 | 0.03172457 |
| R203M | 1.6492504 | 0.15825678 |
| G252V | 1.2301542 | 0.12381144 |
| L452Q | 0.3127277 | 0.03308999 |
| Q498R | 0.2123842 | 0.03172457 |
| N679K | 0.2123842 | 0.03172457 |
| S704L | 0.3077803 | 0.03301503 |
| N856K | -0.2832514 | 0.03621949 |
| Q954H | 0.2123842 | 0.03172457 |
| N969K | 0.2123842 | 0.03172457 |
| L981F | -0.2832514 | 0.03621949 |
| P3395H | 0.2123842 | 0.03172457 |
| I4915T | 0.3189265 | 0.03351622 |
| L5086I | -1.1958547 | 0.13116867 |

**Supplemental Table 1.** **The identified amino acid changes associated estimated effective sizes and standard deviations using the multivariate linear regression model with categorical clusters as covariates.**

| AminoAcidChange | coefficients | Standard deviation |
| --- | --- | --- |
| D3G | -0.5289821 | 0.06197068 |
| T19R | -0.3583990 | 0.05236202 |
| T19I | 0.8675393 | 0.09368981 |
| S26L | -0.3470353 | 0.05215769 |
| T40I | -0.3810594 | 0.05035466 |
| D63G | -0.3508300 | 0.05236169 |
| A67V | -0.5034788 | 0.06039024 |
| I82T | -0.3456858 | 0.05196310 |
| S135R | 0.8074352 | 0.09059436 |
| V213G | 0.2323215 | 0.03223704 |
| T223I | 0.5872878 | 0.08264916 |
| G252V | 1.1976598 | 0.12468634 |
| S371L | -0.5534597 | 0.06193220 |
| T376A | 0.8620845 | 0.08999643 |
| D405N | 0.8797794 | 0.09241162 |
| R408S | 0.6207915 | 0.08287705 |
| S413R | 0.8157651 | 0.09230739 |
| G446S | -0.2664795 | 0.03198609 |
| L452Q | 0.3466409 | 0.03292778 |
| G496S | -0.5499511 | 0.06210286 |
| T547K | -0.5341148 | 0.06147332 |
| P681R | -0.3506782 | 0.05225140 |
| S704L | 0.3418159 | 0.03284982 |
| T842I | 0.7544330 | 0.08850513 |
| N856K | -0.5521347 | 0.06227279 |
| K856R | -0.5393076 | 0.06198051 |
| D950N | -0.3534498 | 0.05196289 |
| L981F | -0.5521347 | 0.06227279 |
| A1306S | -0.3553043 | 0.05035843 |
| G1307S | 0.8259626 | 0.08905459 |
| P2287S | -0.3541119 | 0.05015436 |
| A2710T | -0.5450373 | 0.06223708 |
| V2857A | -1.3198257 | 0.20083279 |
| V2930L | -0.3796890 | 0.05042589 |
| L3027F | 0.8277260 | 0.08812009 |
| T3090I | 0.7634935 | 0.08992720 |
| T3255I | -0.2507549 | 0.03705032 |
| T3646A | -0.3687172 | 0.05037977 |
| I3758V | -0.5466932 | 0.06187469 |
| I4915T | 0.3524137 | 0.03336264 |
| L5086I | -1.2284741 | 0.13159986 |
| S5150F | 0.8465102 | 0.09162000 |
| H5401Y | -0.3689352 | 0.05186462 |
| K6597R | 0.8501061 | 0.09021831 |

**Supplemental Table 2. The identified amino acid changes associated estimated effective sizes and standard deviations using the multivariate linear regression model with MDS-computed dimensions as covariates.**
